# Quantifying intra-individual variations in anatomical sites of pain in longitudinal studies

**DOI:** 10.1101/2025.10.23.25337565

**Authors:** Victoria J Madden, Gwen Arendse, Peter Kamerman, Gillian J Bedwell, Luyanduthando Mqadi, Antonia Wadley, Robert R Edwards, John A Joska, Romy Parker

## Abstract

**Background:** Longitudinal pain studies present an opportunity to understand intra-individual variations in pain. However, current methods to quantify the anatomical extent of pain fail to capture variations across multiple body sites over time. Our aims were to (1) describe patterns of intra-individual variation in pain sites over time, (2) develop a metric that quantifies intra-individual variation in pain sites over time, and (3) determine whether this metric is related to other indicators of pain burden.

**Methods:** We used data from a longitudinal observational cohort of 72 participants (51 female; median (IQR) age 43 (37-51) years) living with virally suppressed HIV, who provided weekly reports of pain severity, pain site(s), and distress over 49 weeks.

**Results:** Participant reports showed noteworthy intra-individual changes in pain sites over time. The pain sites variation metric, based on 727 consecutive reports from 53 participants, reflected intra-individual temporal variation in pain sites, and distinguished participants with consistent pain sites from participants with high temporal variation in pain sites. The metric was positively associated with the count of painful sites, and with emotional distress only when unadjusted for the count of painful sites.

**Conclusions:** Thus, using a longitudinal cohort, we developed a metric that quantifies individual temporal variation in pain sites and demonstrated that it relates more strongly to the count of painful sites than to distress. This metric provides an opportunity to study whether the number of, and variation in, pain sites contribute to pain burden and clinical outcomes.

## Introduction

Repeated measurements of pain reveal high levels of intra-individual variability, whether they are measured in the short term (e.g., during a quantitative sensory testing session) or the long term (e.g., longitudinal clinical trials) (Madden et al., 2021; Mun et al., 2019). While methods for dealing with temporal variability in pain *severity* ratings exist and range from simple (e.g., averaging over repeated measurements (Rolke et al., 2006)) to sophisticated (e.g., calculating indices that incorporate the variability (Mun et al., 2019)), finding suitable methods to represent intra-individual variation in pain *sites* across time remains a challenge.

Recent data from two long-term studies indicate that, even when a person consistently reports pain over time, the site of that pain may vary considerably (Schafer & Zajacova, 2025; Wadley et al., 2021). This variation in pain sites highlights the importance of recording the site(s) of pain in long-term studies, to avoid misinterpreting repeated reports of pain over a prolonged period as persistent pain when they may indicate short-lived pains in multiple sites with temporal overlap.

In cross-sectional studies, pain sites are typically quantified using simple counts. In longitudinal studies, these methods can conceal intra-individual variation over time. Although categorical classifications of change have been used to reduce complexity in research using just two repeated measures (Schafer & Zajacova, 2025), a numerical metric of the temporal variation in pain sites would be more pragmatic for studies using more repeated measures. Tracking absolute pain site counts at each sampling interval, or changes in the count of endorsed sites (e.g. Erickson et al., 2021), is inadequate. For example, a person who reports head pain in week 1, and pain in their hip, ankle, and shoulder (but not their head) in week 2, the pain site count has gone from 1 site to 3 sites, a change of 2 sites—but the person has actually reported 4 changes in pain sites because a negative change (loss of the head as a pain site) may be as relevant as a positive change (gain of hip, ankle, and shoulder as pain sites). A metric of temporal variations in body site endorsement must capture changes in site, regardless of the direction of change.

Here, we report our approach to describing and quantifying variation in weekly measurements of pain sites. We use data from a primary study that investigated the effect of psychological distress on pain in people living with HIV (Madden et al., 2022). We consider this cohort ideal for studying pain site variation because people living with HIV have previously been found to have high variation in pain sites (Wadley et al., 2021). The current dataset provides fine-grained data on week-to-week fluctuations in sites and severity of pain. In addition to describing patterns of intra-individual variation in pain sites and developing a metric that quantifies that variation, a secondary objective was to determine whether our pain sites variation metric was positively associated with the within-participant mean count of painful sites, pain severity, or variance in pain severity, or emotional distress.

## Methods

### Study overview

This longitudinal, observational study entailed weekly repeated assessments of emotional distress and pain over six months. Participants self-reported distress and pain via a mobile phone app (Upinion, Opinion Solutions (Pty) Ltd) using their personal or study-provided mobile phone (Mobicel FAME 16GB). The question battery was available in either English or isiXhosa (participant’s choice). Current distress was assessed first, followed by questions on pain in the past week (reported here) and for the past three months (not reported here). Airtime compensation and a raffle system were used to incentivise participants to complete the weekly question battery. Upon weekly review of responses, non-responding participants were contacted telephonically to offer necessary technical support; participants reporting consistently high distress were offered information on local mental health services.

Approval for this study was obtained by the Human Research Ethics Committee of the University of Cape Town (approval number: 764/2019) and the City of Cape Town (ref: 24699). We followed the STROBE reporting guidelines (Vandenbroucke et al., 2007) (Supplementary file). Details of the current study procedures are listed in a protocol locked online (Madden et al., 2022). Withdrawal from the study was permitted with no repercussions. Participants and data collection staff were not told about the study hypotheses; no blinding assessments were conducted.

### Procedure

#### Participants

The cohort for the current study was drawn from a primary study that had aimed to include a pragmatic sample of 100 consenting adults with suppressed HIV who had consistently reported either persistent pain or no pain across the screening and baseline assessments for that study, and had no indication of psychiatric problems (see (Madden et al., 2022) for details). We invited all those people to opt into this cohort with no additional eligibility criteria, regardless of baseline pain status.

### Outcomes

#### Pain

Pain-related questions were modified from the Brief Pain Inventory (BPI) (Parker et al., 2016). Questions elicited information on pain presence/absence, anatomical site(s) of pain, anatomical site of the worst pain, and average and worst pain severity at the site of worst pain (Figure 1). Here, we use the questions asked about *pain in the past week*. Pain sites were reported on a body map divided into 18 regions, a similar approach to the Michigan body map (Clemens et al., 2024). Anatomical site name, colour, and number indicated the regions (Figure 2a). Pain severity (at the site of worst pain) was rated by selecting a number from 0 (no pain) to 10 (pain as bad as I can imagine) on a vertical visual & numerical scale (Figure 2b).

**Figure 1.**
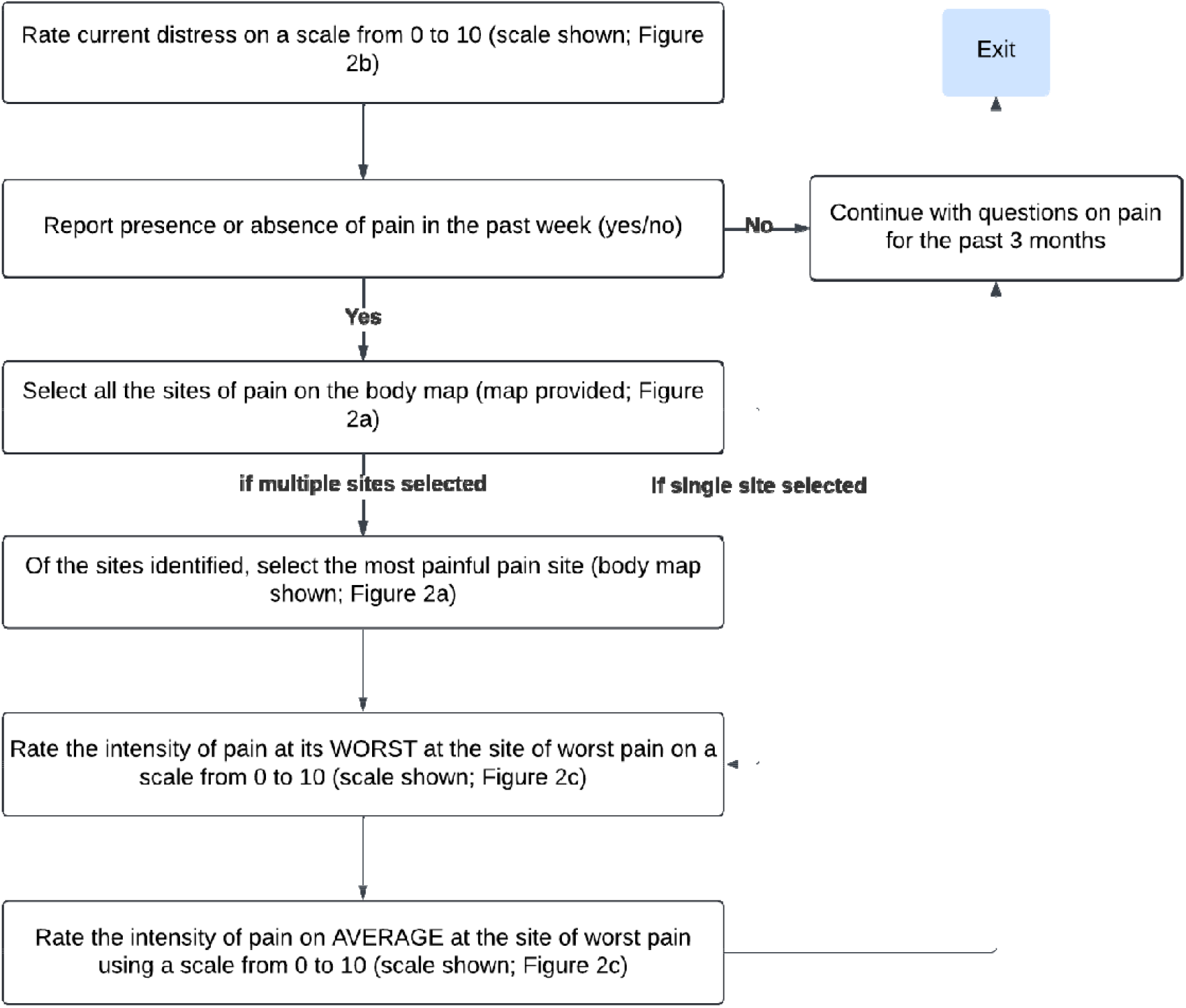
Question flow. Exact phrasing is shown in Figure S1.

**Figure 2.**
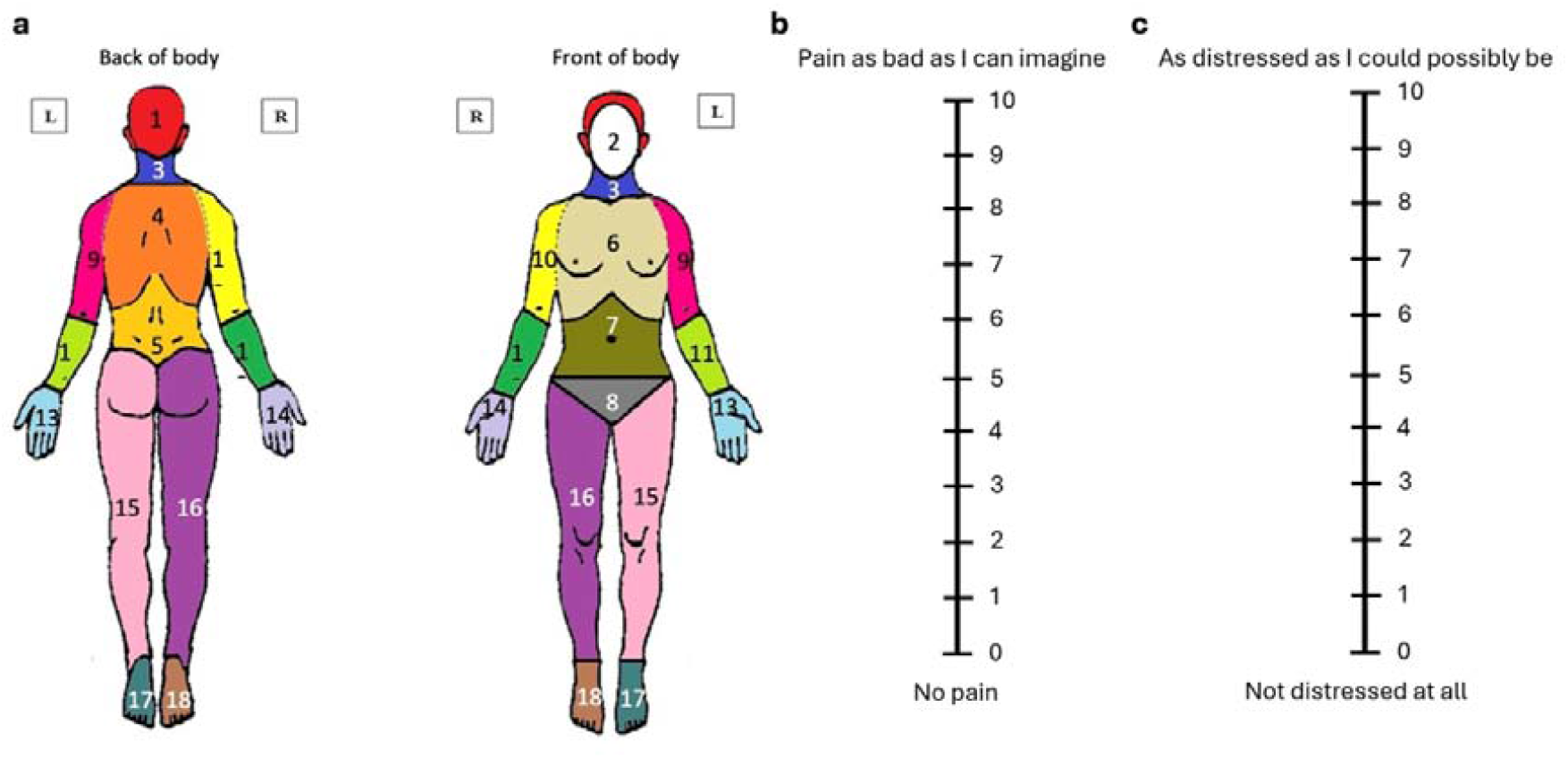
a) Body map, b) visual & numerical scale for pain severity and c) visual & numerical scale for distress.

#### Emotional distress

Emotional distress was reported on a vertical visual & numerical scale in response to the question, “Throughout our lives, most of us feel distressed from time to time. Select the number between 0 and 10 that best describes how distressed you have felt, on average, over the past week.” The scale ranged from 0-10, with 0 at the bottom of the scale indicating ‘not distressed at all’ and 10 at the top, indicating ‘as distressed as I could possibly be’. Participants could select any number from 0-10 (Figure 2c). The within-participant mean of distress scores (sum of distress scores divided by the count of distress scores) was taken forward to analysis.

### Data analysis

This exploratory analysis visualised and statistically analysed distress and pain data reported over multiple weeks. The data collected on the mobile phone app were captured on Excel spreadsheets, weekly, with the date of assessment. The data were imported from the Excel spreadsheets and combined into a full dataset in R (R Core Team, 2023) (via RStudio (Posit team, 2025)).

#### Data review and cleaning

The study ran for 49 weeks; each response was labelled with a number denoting the week in which that response was given. Week 1 began on the 16th of March 2021.

Data were excluded from seven participants due to data being inconsistent with eligibility criteria for the primary study (n = 6 screening and initial data collection questionnaires showed conflicting information on pain status, suggesting unreliable responding; and n = 1 participant was pregnant). We identified a further seven participants who had completed the survey more than once in one week. These responses were assessed for credibility. First, we compared the participant study identifier (PID) reported by the participant to the PID that had been assigned to the app-generated user number at study enrolment. Second, we assessed the consistency of responses across the weeks that preceded, included, and followed the week with multiple responses. Where there was clear consistency, the responses were flagged as duplicates, and the duplicate copy was removed (n = 4 responses). Inconsistent responses underwent logic checks, and subjective judgements were used to establish whether distress or pain responses were realistic. Again, responses were removed in cases with reasonable doubt (n = 3 responses).

#### Descriptive statistics and plots

The median and interquartile range (IQR) were used to describe numerical data; frequency and proportion describe categorical variables. We descriptively and visually report the presence of pain, sites of pain and pain severity at the individual and group levels. Given the low response rate in the sample, we used linear regression models to assess whether missing responses were associated with participants’ age, sex, mean distress rating, mean of the worst pain intensity, or the mean of the count of painful sites.

#### Statistical analysis

The goal of the current analysis was to develop a metric to capture variation in pain sites, after data inspection revealed the need for such a metric. The metric was intended to represent the total number of week-to-week changes in pain sites relative to the total number of week-to-week reports provided by the participant. Therefore, we included data from participants who had endorsed having pain at least once and, at a minimum, provided a response to the first question about pain in two consecutive weeks, during the study. We then proceeded using only those responses that were temporally consecutive to another response, within that participant (i.e. had a response in the immediately preceding week), and that therefore provided an opportunity to capture potential change in pain sites. In the pain sites variation metric, the numerator is the sum of all changes in endorsed pain sites between each pair of consecutive responses across the study. The denominator is the number of opportunities to observe change—that is, the number of consecutive response pairs provided by the participant. The first response in any sequence is not included in the denominator, because a single response alone cannot show change. The pain sites variation metric for a participant is defined as:

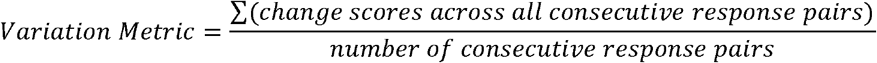

For example, if a participant reported pain in weeks 1, 2, and 5, only the change from week 1 to week 2 contributed to the metric; week 5 did not, because it was not preceded by a consecutive report (see Table 1). If a participant endorsed chest pain only in one week and the next endorsed head pain, but not chest pain, this represented two site changes and contributed a change score of 2 to the numerator. That pair of reports contributed 1 to the denominator (e.g. PID X1 in Table 1). An adaptable script to calculate the metric is available at [https://osf.io/xuvjg/overview?view_only=4f4375b56f764d6c86c616744a313989].

**Table 1.**
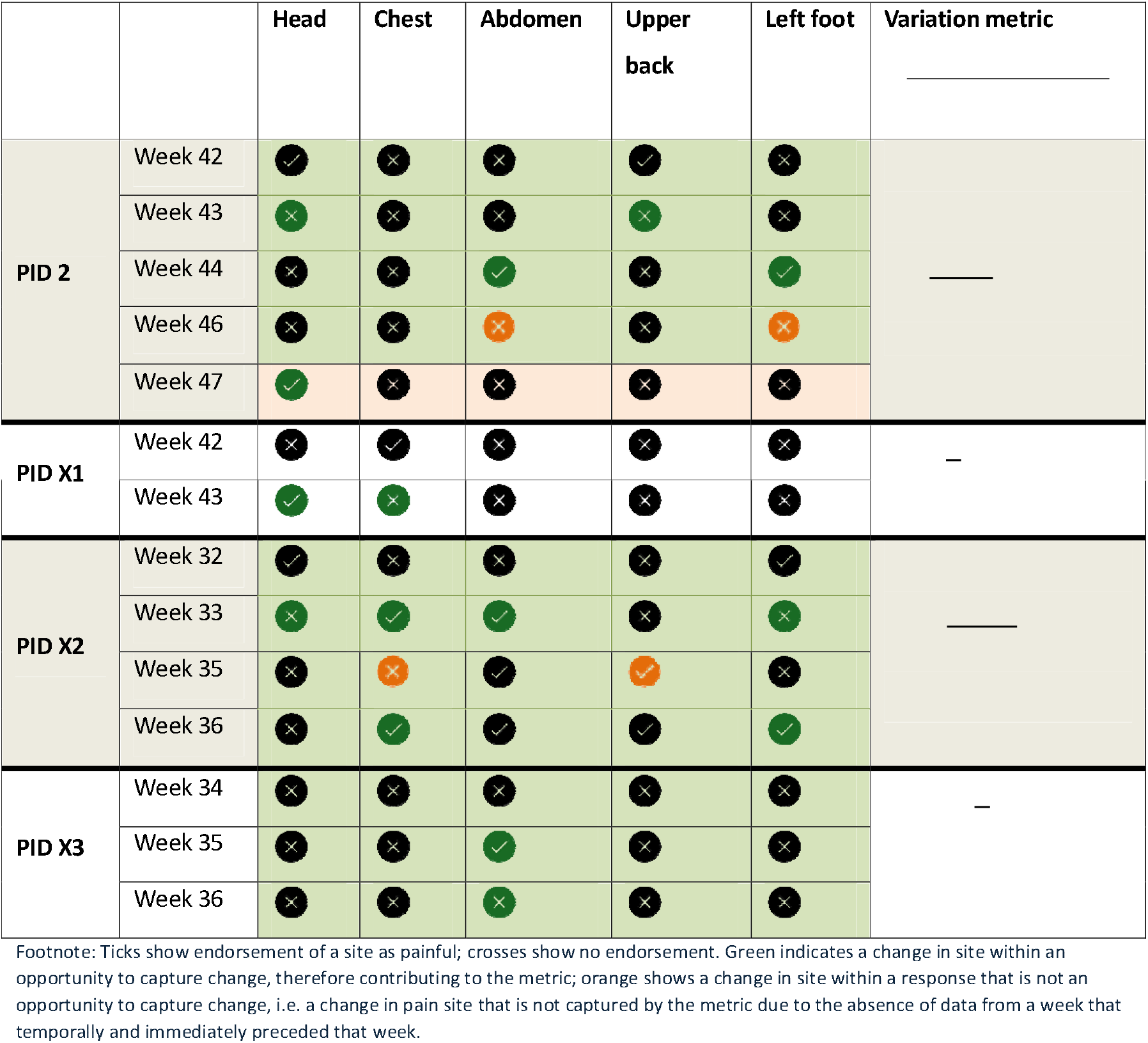
Examples responses from real and dummy participants (dummy PIDs indicated with X) to illustrate changes in pain sites from the preceding week and how the variation metric is calculated to reflect changes divided by opportunities to capture change.

First, we compared the metric with visualised individual endorsements of pain sites over time. In the specific context of the pain site variation metrics found in the current sample, we labelled variation as comparatively low, moderate, or high. Importantly, these descriptive labels are specific to the current sample and not intended to be transferable to other samples or studies. Second, we assessed the range of the metric by plotting its distribution at the sample level, with four different cutoffs for participant inclusion. The purpose of this was to understand the metric’s potential usefulness for studies with different numbers of repeated assessments. The cutoffs were informed by the data available; we used participants who had provided data with at least 1, 6, 11, or 21 opportunities to capture change. Third, we assessed the relationships between the pain sites variation metric and participant sex using an independent t-test. Fourth, we used Spearman’s rank’s correlation test to assess whether the pain sites variation metric was associated with the mean of the count of painful sites (because more sites of pain should support more variation), the mean of worst pain severity, the standard deviation of worst pain severity (as a measure of variance in pain severity), or mean distress. We report the correlation coefficients (rho) and interpret p-values with an alpha of 0.05. We also performed a linear regression to assess whether associations shown to be significant with the tests of simple correlation remained so when adjusted for the mean of the count of painful sites. We report the estimates with 95% confidence intervals and p-values at a threshold of 0.05. Fifth, to test whether these correlations differed systematically between people who contributed data with different numbers of opportunities to capture change, we conducted a weighted correlational analysis by assigning the data from each individual a weight according to the total number of opportunities to capture change in their data (e.g. the top contributor of 29 opportunities to capture change received a weight of 29/29 = 1; the lowest contributor of 1 opportunity to capture change received a weight of 1/29 = 0.034). We then compared the correlation coefficients between the weighted and unweighted analyses.

This analysis was conducted in R version 4.4.0 (R Foundation, Vienna, Austria) and RStudio (version 2024-04-24), using packages knitr (Xie, 2024), dplyr (H. Wickham, & François, R, 2023), tidyverse (Wickham et al., 2019), gridExtra (Auguie, 2017), magrittr (Bache, 2022), gtsummary (Sjoberg, 2021), flextable (Gohel, 2024), ggplot2 (Wickham, 2024), patchwork (Pedersen, 2024), data.table (Barrett et al., 2024), lubridate (Grolemund, 2011), forcats (H. Wickham, 2023), formatR (Xie, 2023), glue (Hester & Bryan, 2024), sjPlot (Lüdecke D, 2024), purrr (Henry, 2024), weights (Pasek et al., 2025), rlang (Henry & Wickham, 2026), here (Müller & Bryan, 2025), webshot (Chang, 2023), wCorr (Bailey & Emad, 2023), and car (Fox & Weisberg, 2019). All participant data were de-identified for this analysis; the PIDs presented in the plots are different from the ones used during the data collection process.

## Results

### Description of all data

The study ran for 49 weeks and enrolled 72 adults. Most respondents were female (n = 51; 71%) and the median age (IQR) was 43 (37-51) years. In total, we received 1070 responses from these 72 participants. The number of responses provided by each participant ranged from 1-40. Only one participant provided 40 responses; the median (IQR) number of responses was 10 (5-17). A greater number of missing responses was associated with being male, reporting greater distress, and reporting more severe pain (Table S1). Due to variation in individuals’ contributions to the overall dataset, we do not present participant-reported distress or pain variables at the level of the whole dataset because variable contributions per participant yield biased group-level data. Overall, 53 participants provided 2 or more consecutive responses including pain site data, across 49 weeks. That is, 53 participants provided ‘opportunities to capture change’ that were carried forward to the calculation of the pain sites variation metric. Of these 53 participants, three had a mean distress rating of 10/10. The mean rating of worst pain was greater than 3/10 for most of the 53 participants; the most common mean rating of worst pain was 9/10 (n=9 of 53 participants) (Figure 3a). Across the opportunities to capture change in pain sites (i.e. the data used to calculate the pain sites variation metric), the most common within-participant mean distress rating was 6 or 8/10 (n = 7 each), followed by 4 (n = 6) (Figure 3b).

**Figure 3.**
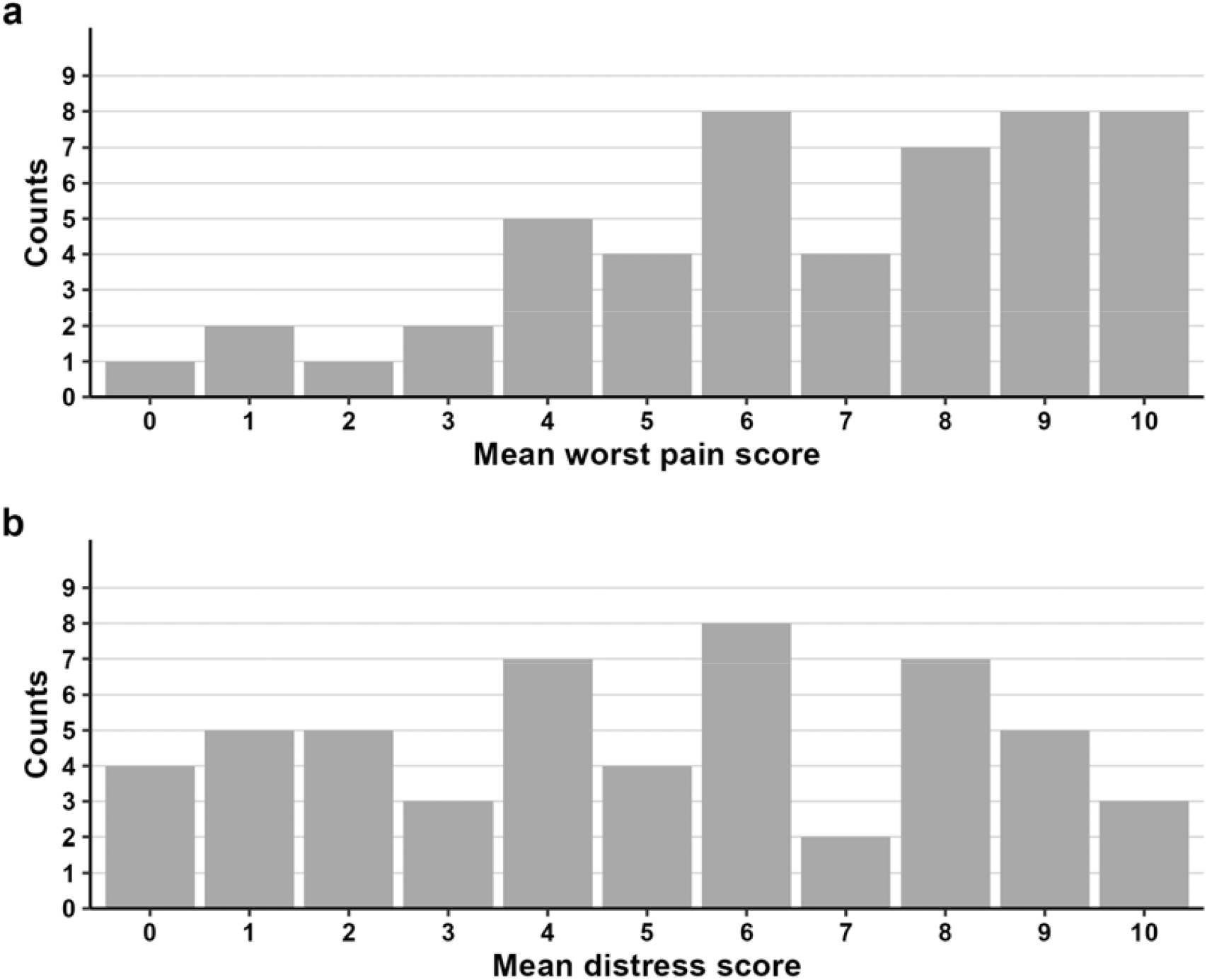
a) Mean worst pain severity and b) mean distress scores, for each participant; usable responses only.

### Variation in the presence of pain

Figure 4 shows the opportunities to capture change provided by each participant over time (n=53). The median (IQR) number of opportunities to capture change provided by a participant over the course of the study was 10 (5-17). Over the course of the study, 5 participants provided the minimum of 1 opportunity to respond (PIDs 9, 20, 21, 43 & 47); 2 participants (PIDs 27 & 34) provided the highest number of 29 opportunities to capture change, both covering a continuous 30 week-period. Although all participants had been enrolled to the parent study after reporting either no pain or persistent pain, in these weekly remote responses, endorsement of the presence of pain across the opportunities to capture change was mostly inconsistent (note that data from a participant *never* endorsing pain would have been omitted for providing no opportunity to capture change). Only three participants (PIDs: 7, 34 & 41) reported pain in every consecutive week after their first response to the question. Nine participants (PIDs: 3, 18, 21, 22, 25, 29, 40, 47 & 53) were consistent in their endorsement of pain whenever they did respond, but did not respond in every week after first responding. The remaining n = 42 had inconsistent pain status responses.

**Figure 4.**
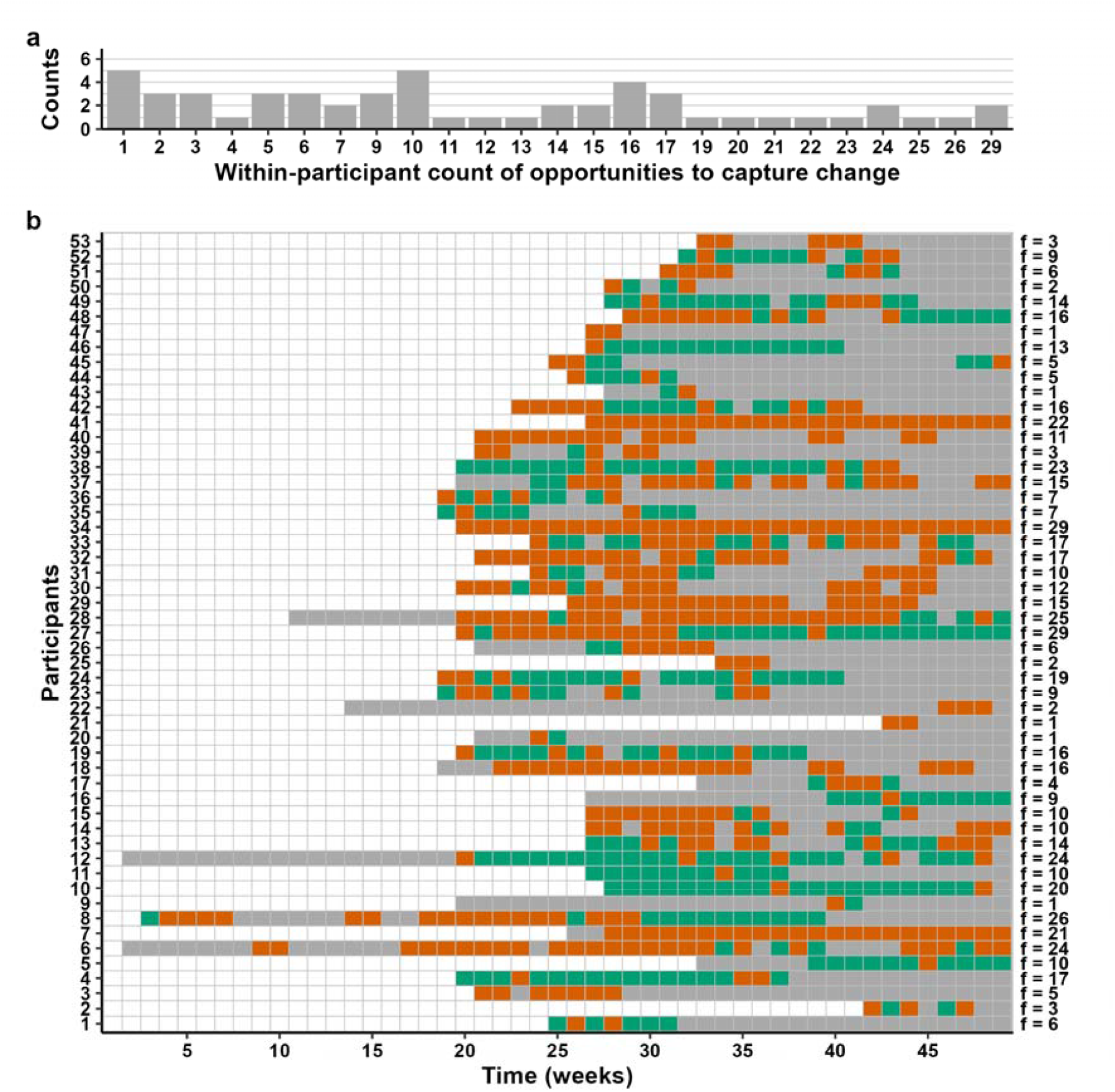
Frequency of a) opportunities to capture change and b) presence of pain in the past week across 49 weeks. Orange colour: participant endorsed *pain* in the last week; green colour: participant endorsed *no pain* in the last week; white colour: weeks before a participant was enrolled into the study; grey colour: participant failed to respond *or* week without a response in the immediately preceding week). Study mobile phones were distributed to participants from 20 weeks into the study, to support participation. The number of opportunities to capture change for each participant is indicated on the far right with f (denoting frequency).

#### Pain sites

We saw marked intra-individual variations in pain sites over 49 weeks. From the 53 participants who provided data with opportunities to capture change, Figure 5a shows the count of endorsements of each possible number of unique pain sites (maximum 18) reported by each participant over the course of the study. It was most common for participants to endorse only two pain sites across the 49 weeks (n = 9), although five participants endorsed one pain site, and one participant endorsed every one of the possible 18 pain sites at some time during the study. Figure 5b shows endorsement rates for each of the 18 pain sites, at the group level - with endorsement of a site counted only once for each participant (n=53). The pain sites most commonly endorsed at least once were the head, upper back, lower back, upper arm, feet, pelvis, and forearms (Figure 5b).

**Figure 5.**
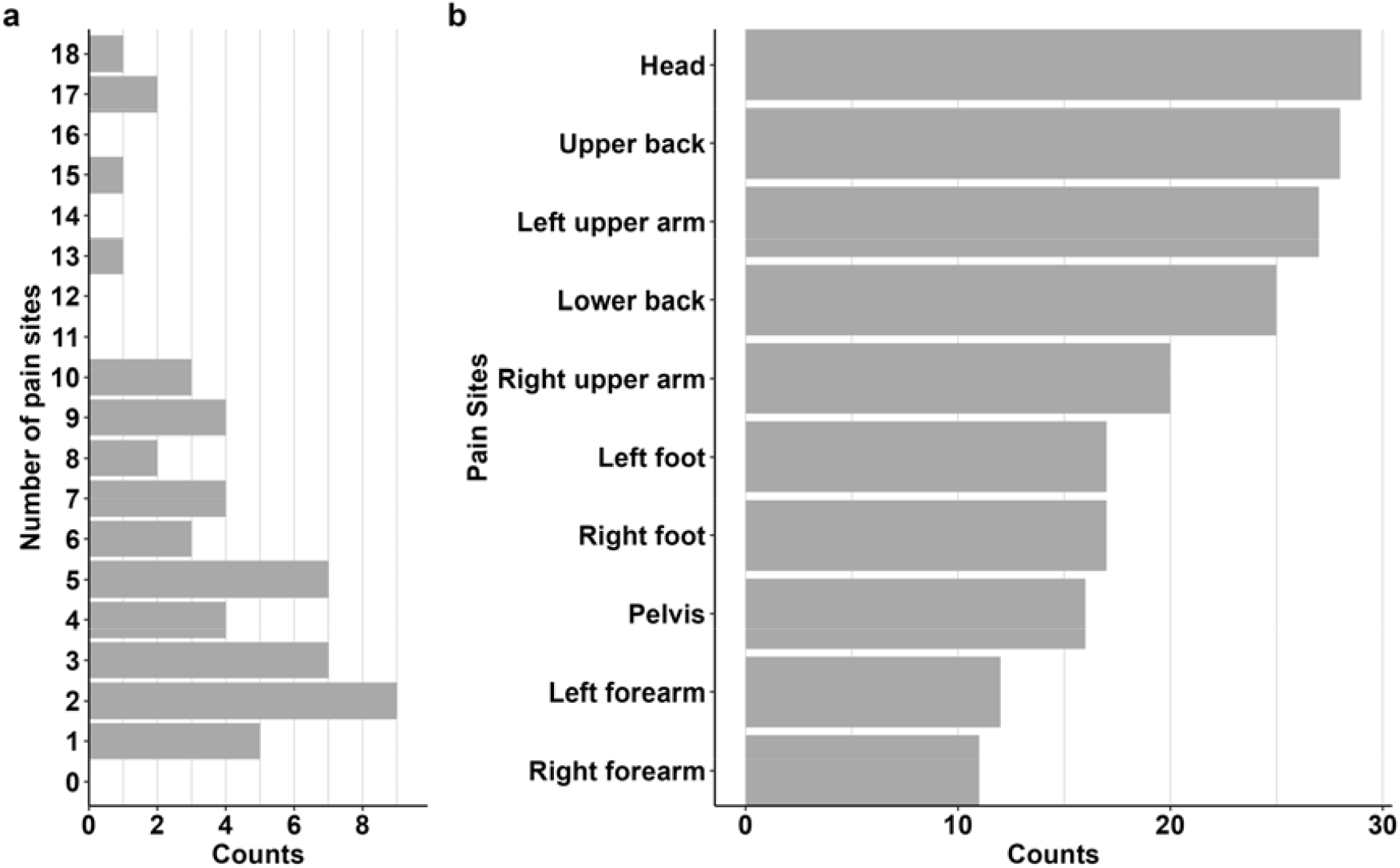
For all participants who provided data with opportunities to capture change, pain in the past week: a) count of endorsements of each possible number of pain sites b) the count of times each site was endorsed at least once by a participant. Numbers indicate endorsement counts.

#### Pain sites variation metric

The 53 participants who provided data with opportunities to capture change together provided 727 responses over the course of the study. Figures S2-S19 show the pain sites endorsed by each participant over time, along with their variation metric; Figure 6 shows three of these plots selected to represent participants with low, intermediate, and high variation in pain sites, relative to the current sample. Of the 53 participants, 5 participants reported a single site of pain; these participants and those who reported minimal week-to-week variation in pain sites had relatively low variation metrics (e.g. Figure 6a, PID 5). In contrast, 39 participants (e.g., PIDs: 14, 28) reported more than one site of pain at least once. Participants who frequently reported different sites, including over consecutive weeks, had relatively higher variation metrics (e.g., Figure 6b & c). PID 28, who reported multiple, varying pain sites over time, had the highest variation metric in the sample (Figure 6c, variation metric = 5.11).

**Figure 6.**
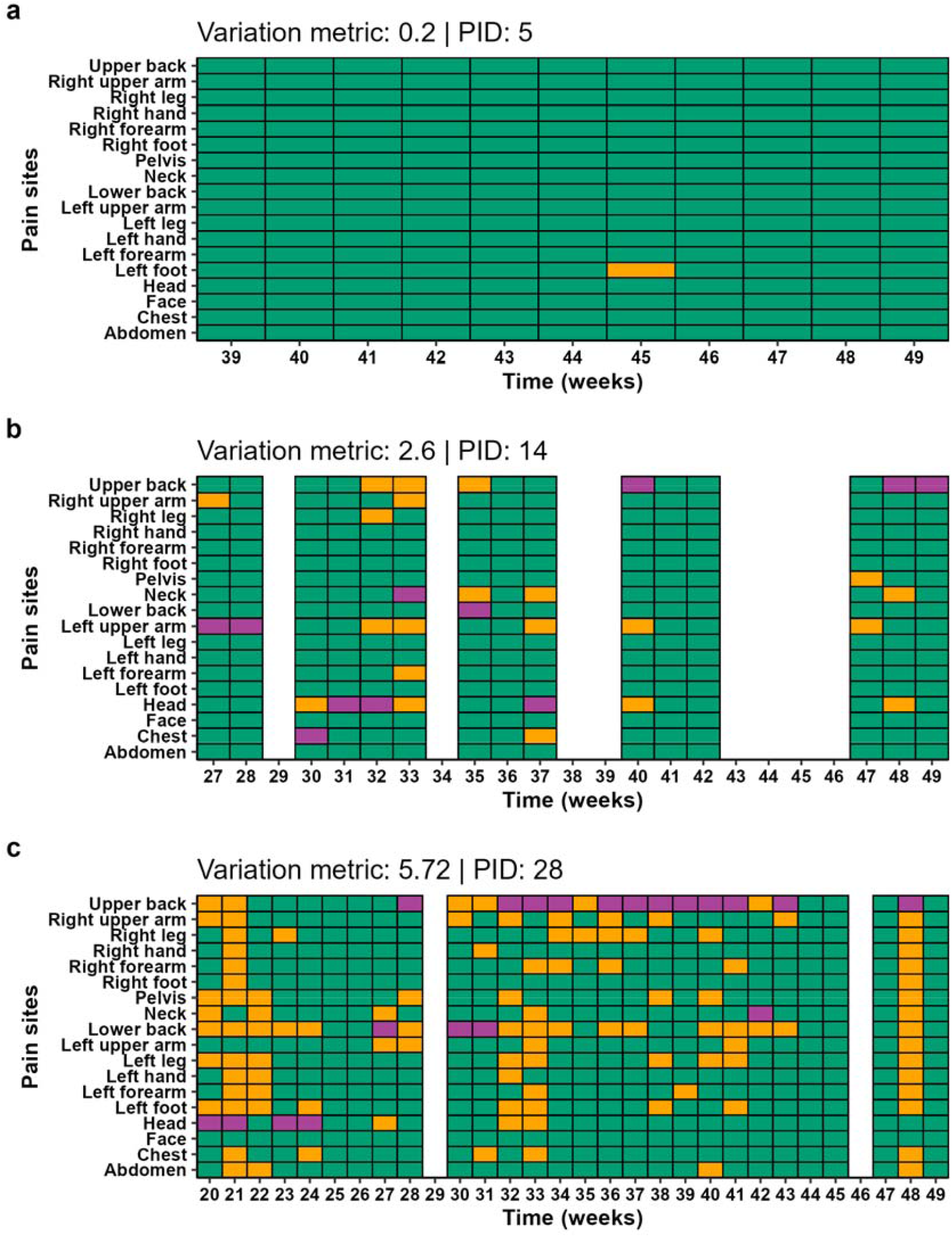
Sites and severity of pain in the past week for 3 individuals (a-c). Each orange block shows an endorsed pain site, and the worst pain site is presented by a purple block; no pain site endorsed is indicated with a green block. Weeks with missing responses are shown in solid white.

Figure 7 shows the distributions of the pain sites variation metric based on the four different cutoffs for participant inclusion in this analytical step, which were greater than or equal to 1, 6, 11, or 21 opportunities to capture change. The range of the pain sites variation metric was consistent for the four cutoffs, at 0.18 to 5.11 (Figure 7a-d), but there was some difference in the distribution of the metric: the median (IQR) variation metric was higher for ≥21 week of responses (1.18 (0.42-1.92)) and lower for ≥11 weeks of responses (1.06 (0.45-1.92)). Few participants had a variation metric greater than 3 (n = 5 for ≥1 week; n = 4 for ≥6 weeks; n = 3 for ≥11 weeks and n = 1 for ≥21 weeks).

**Figure 7.**
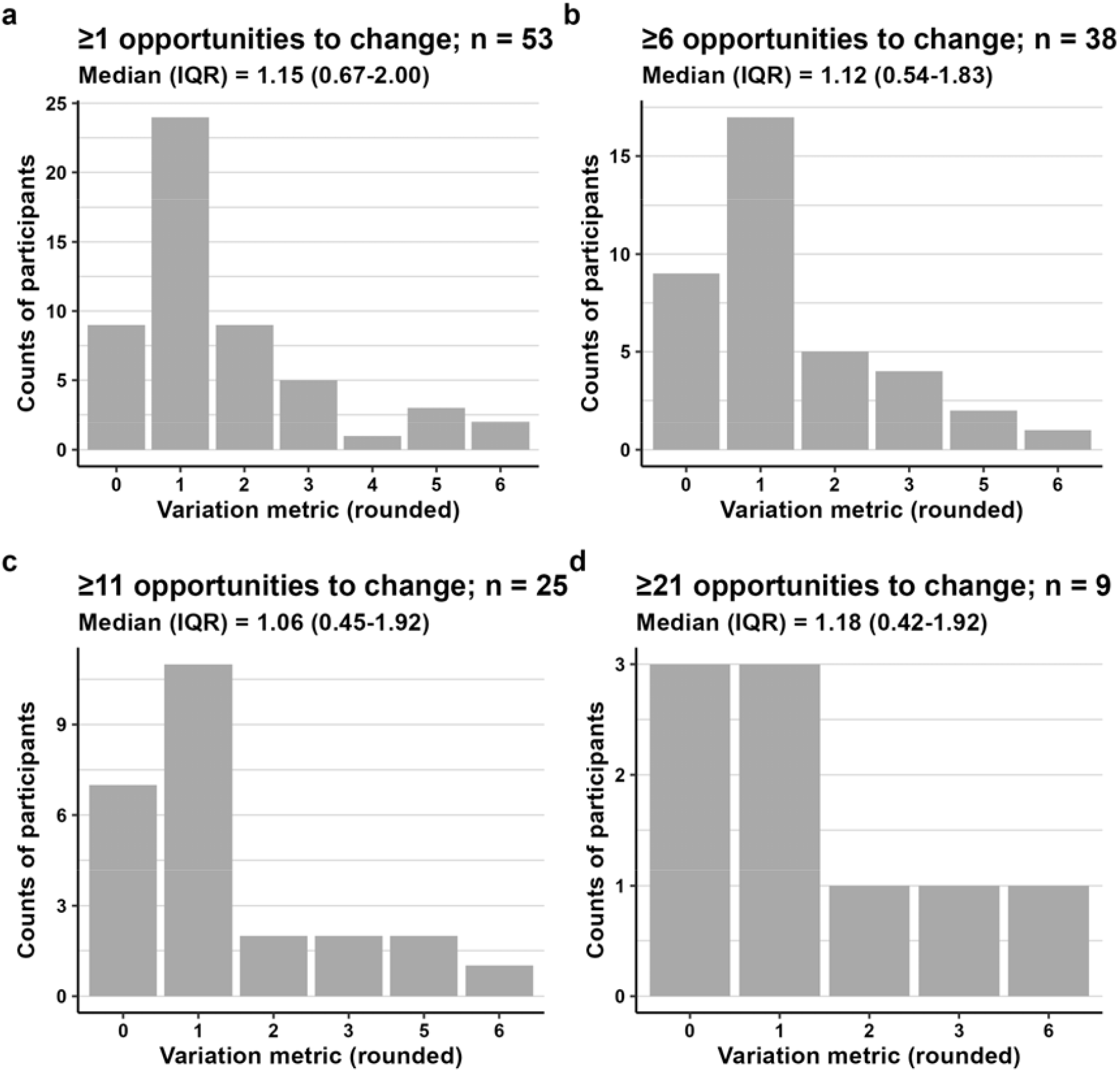
Variation metrics for subsample defined by having provided data with opportunities to change (a) ≥1, (b) ≥6, (c) ≥11, or (d) ≥21 weeks.

### Relationships of the metric to demographic and clinical features

The value of the pain sites variation metric was not associated with participant sex (mean in males = 1.49, mean in females = 1.66; p = 0.127). Table 2 shows the Spearman’s correlation test results from the unweighted and weighted analyses, at each subsample cutoff, for the relationships between the pain site variation metric and the within-participant mean of the count of painful sites, the within-participant mean of the worst pain severity, the standard deviation of worst pain severity, or mean distress (also see Figures S19-S23). The pain sites variation metric was strongly correlated with the mean of the count of painful sites at all cut-offs except at ≥21 opportunities for change in the unweighted Spearman’s rank correlation analysis. There was no evidence that the metric was associated with the mean nor the standard deviation of severity of the worst pain (Table 2). The pain sites variation metric was positively and moderately associated with distress at all cutoffs except ≥21 weeks, where the correlation coefficient remained moderate. This association was also no longer significant once adjusted for the mean count of painful sites (Table 3).

**Table 2.** Spearman’s Correlations between the pain sites variation metric and the within-participant (a) mean of the count of painful sites, (b) mean of ratings of worst pain severity, (c) standard deviation of ratings of worst pain severity, or (d) distress ratings, using Spearman’s correlation test and unweighted and weighted analyses, at each subsample cutoff. The asterisk (*) denotes a statistically significant correlation at alpha=0.05 (uncorrected).

|  | Spearman's rho |  |
| --- | --- | --- |
|  | unweighted analysis | weighted analysis |
| d) For mean of the count of painful sites |  |  |
| ≥1 opportunities to change (n=53) | 0.83* | 0.73 |
| ≥6 opportunities to change (n=42) | 0.78* | 0.72 |
| ≥11 opportunities to change (n=32) | 0.75* | 0.68 |
| ≥21 opportunities to change (n=12) | 0.48 | 0.42 |
| d) For mean of ratings of worst pain severity |  |  |
| ≥1 opportunities to change (n=53) | 0.19 | 0.10 |
| ≥6 opportunities to change (n=42) | 0.21 | 0.088 |
| ≥11 opportunities to change (n=32) | 0.11 | 0.025 |
| ≥21 opportunities to change (n=12) | -0.49 | -0.51 |
| d) For standard deviation of ratings of worst pain severity |  |  |
| ≥1 opportunities to change (n=53) | 0.014 | 0.12 |
| ≥6 opportunities to change (n=42) | 0.089 | 0.14 |
| ≥11 opportunities to change (n=32) | 0.067 | 0.13 |
| ≥21 opportunities to change (n=12) | 0.44 | 0.47 |
| d) For mean of ratings of distress |  |  |
| ≥1 opportunities to change (n=53) | 0.42* | 0.45 |
| ≥6 opportunities to change (n=42) | 0.58* | 0.49 |
| ≥11 opportunities to change (n=32) | 0.52* | 0.44 |
| ≥21 opportunities to change (n=12) | 0.067 | -0.0090 |

**Table 4.**
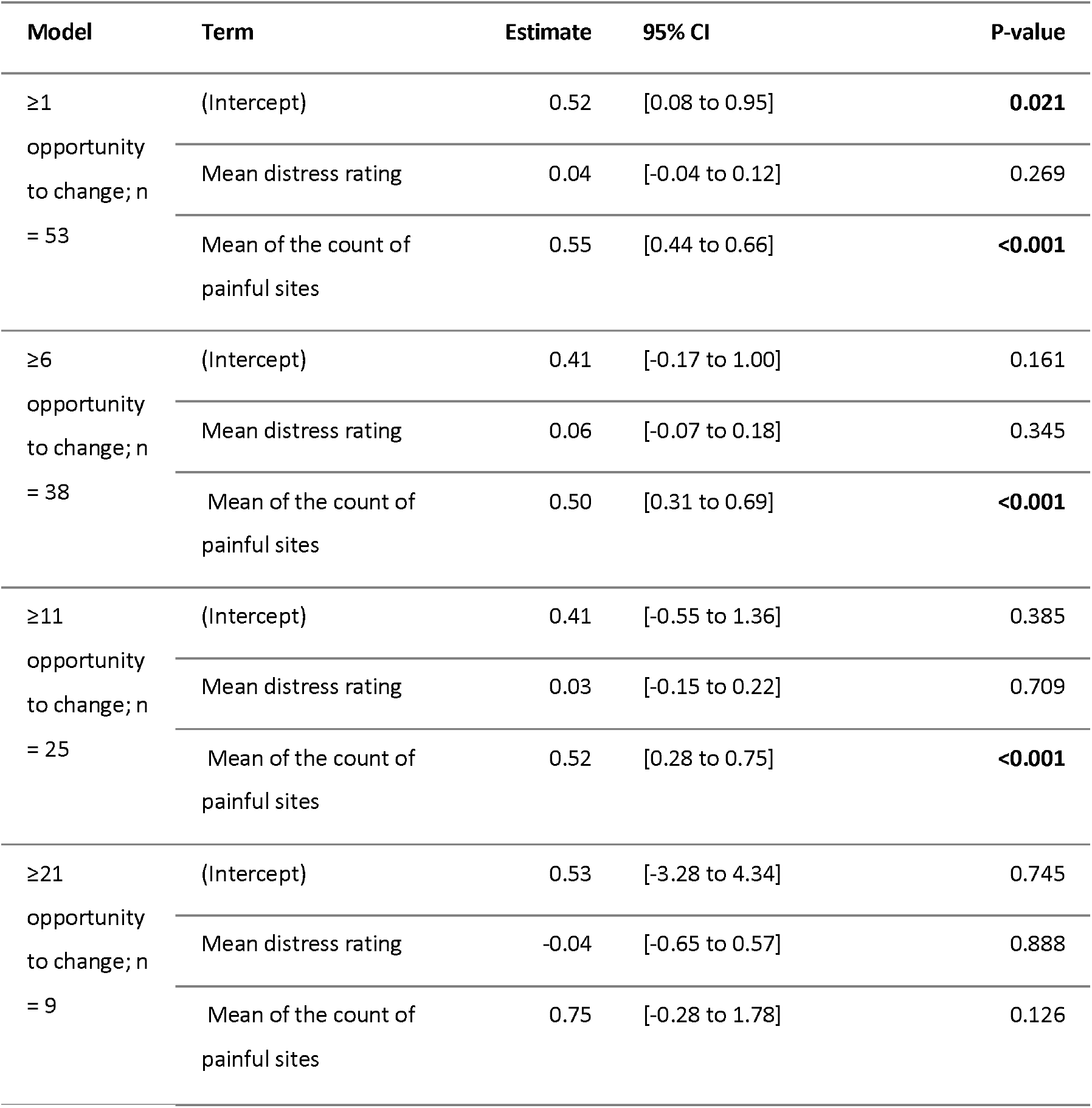
Linear regression of distress predicting the pain sites variation metric adjusted for the mean of the count of painful sites. Significant results at p <0.05 are shown in bold. CI = confidence interval.

## Discussion

This study developed a metric to describe and quantify intra-individual variation in pain sites for human longitudinal pain studies, and determined whether the metric was associated with pain extent, pain severity, variance in pain severity, or emotional distress. We drew on data from a cohort of 53 participants who, together, had provided 727 weekly reports of pain severity, pain site(s), and distress over 49 weeks, and reported marked intra-individual changes in pain sites and pain severity between responses. The pain sites variation metric successfully captured the visualised intra-individual temporal variation in anatomical sites of pain by distinguishing participants with more consistent pain sites from participants with higher temporal variation in pain sites. The metric showed reasonable spread across the cohort. The pain sites variation metric was positively associated with the count of painful sites in unweighted analyses for 3 of the 4 inclusion cutoffs, suggesting, as expected, that people with more pain sites also had more variation in those sites. The metric was also associated with emotional distress in unweighted analyses for 3 of the 4 inclusion cutoffs, but this relationship did not remain when adjusted for the count of painful sites – suggesting that distress may relate more closely to the number of painful sites than to variation in painful sites. Interestingly, that these relationships were not observed in the weighted analysis suggests that they may be more prominent in those participants who provided more data. Perhaps participants whose pain distribution is linked to emotional distress are more motivated to respond to questions about their pain, although data missingness was also more common amongst those with more severe pain and more distress.

Variation in pain severity has recently received increased attention (Edwards et al., 2016; Mun et al., 2019), yet published studies of variation in pain *sites* remain uncommon—and our analysis suggests that variation in severity and sites may not track together. A study of chronic pelvic pain assessed pain sites 4 times a day, and represented intra-individual variation in pain sites using the within-individual standard deviation in the count of pain sites over the 14-day study period (Erickson et al., 2021). However, since this approach cannot capture both positive and negative changes in pain sites, it may under-represent participants’ lived experiences of change in pain sites. Another study, in older Americans, categorised pain site changes between two study assessments as ‘stable, spread, switch, or subside’ (Schafer & Zajacova, 2025)—which nicely represents real-life changes in pain sites, but only across two repeated measures. While we argue that a better metric should capture variation across repeated measures of pain sites, these studies did note intra-individual variation in pain sites that demands focused research.

The significance of intra-individual variation in pain sites is not yet known. In our data, people with more sites of pain also had more variation in those pain sites. Similar to variation in pain severity and to the count of painful sites, variation in pain sites may be clinically meaningful as an outcome or a predictor. For example, is higher variation in pain sites associated with greater impact of having pain, in people with chronic pain? Our analysis suggests pain site variation may not independently predict emotional distress; rather, our findings resemble other evidence that people with more sites of pain report more distress (Iyar et al., 2019; McMillan et al., 2010; van der Windt et al., 2002). However, it remains possible that people living with pain do better when the bodily locations of their pain are more consistent; this is an outstanding question for direct engagement with people living with pain.

Given the priority that many people with pain place on pain controllability and predictability, and the extra resources required to adjust to frequent changes in circumstances (Nichols et al., 2024), reducing variation in pain sites may be important to people living with pain. However, we caution that most current work on pain controllability and predictability has implicitly focused on pain severity or interference, leaving a current shortage of previous research on pain *sites* to ground this idea.

Variation in pain sites seems likely to influence other clinical outcomes, given that changes in pain sites are difficult to interpret and difficult to manage. We have observed that clinicians and patients have difficulty making sense of ‘pain that moves’, complicating diagnosis, treatment, and both psychological and functional coping. It would be worthwhile to directly investigate whether variation in pain sites predicts non-pain outcomes including psychological processes (e.g. pain-related worry or fear, hopelessness), mental wellbeing, functional goal attainment, and quality of life. The relationship of pain site variation to treatment responsiveness may also be worthy of attention. In clinical trials, higher variation in pain severity at the baseline time point has been associated with greater response to placebo interventions (Scott-Lennox et al., 2001) and smaller response to active pharmacological interventions (Mun et al., 2019), suggesting that higher variation in pain severity could serve as a variable for enriching trial cohorts. Crucially, variation in pain sites has not received enough attention to clarify whether it, too, could predict responses to treatment. The metric we offer here should facilitate such inquiry: it is simple to calculate without specialised software, and we have provided a simple, adaptable R script to facilitate this.

In the clinical context, variation in the sites of a person’s pain often informs diagnostic and treatment decisions. Similarly, in the research context, pain site variation data has the potential to inform tentative deductions about putative causes of pain. For example, if a person reports bilateral foot pain but only feels it intermittently across consecutive weeks, we could reasonably rule out distal sensory neuropathy as a dominant cause. Similarly, if a person reports pain in one location and no pain in that location the following week, physical injury seems an unlikely cause of that pain. In this way, information on intra-individual variation in pain sites could support a degree of deductive reasoning that would be useful in interpreting data from large study cohorts where full diagnostic work-up is unavailable.

The data from this cohort were reported remotely, which relies on participant responsiveness and offers limited opportunities for data validation. To support responses and identify misleading data, we frequently offered participants training and app-use support, called non-responders, checked on those reporting high distress or pain, and referred distressed participants to community services. To detect potential proxy reporting, we cross-checked each participant-reported confidential study identification number against the number entered by the research team at app installation. Several participants commented on the intuitive nature of the chat-like app interface. These strategies and feedback give us confidence that our data represent remote reporting on pain and distress from our participants’ real-life context of daily living. A second consideration is that the list of pain sites began with the head (Figure 2). It is possible that the high endorsement of the head as a pain site was biased by this presentation, however, the reasonably high endorsement of the feet (bottom of the list) suggests that participants accessed the entire list.

In this work, we have proposed that, alongside intra-individual variation in both pain severity and interference, intra-individual variation in pain sites may be an important measure of pain. We have provided a single metric to represent intra-individual variation in pain sites over time, capturing both positive and negative changes at each bodily site and the frequency of those changes over time. An important limitation of the metric is its dependence on the number of anatomical sites into which the body is divided for the reporting task. Our participants could endorse up to 18 sites; a different number of possible sites would yield different numbers for the metric. This limits external comparison of the values of the metric across studies with different methods. The values of the pain sites variation metric in the current dataset are also not suitable for use as normative values. However, the metric seems well suited to within-study use to understand the relevance of variation in pain sites for wellbeing, given that our analysis was able to demonstrate that its relationship to emotional distress was removed by adjusting for the mean count of painful sites.

The next challenges in understanding the importance of variation in pain sites are to investigate the extent to which pain site variation matters to people living with pain, and whether it belongs in a causal chain modulating other outcomes or is a consequence of changes in other outcomes. Achieving this clarity will likely require intensive longitudinal study designs that provide high temporal resolution across multiple measures of pain and related factors such as psychological wellbeing, social interactions, goal attainment, and healthcare experiences. Given the complex, interconnected nature of pain, it will be important to analyse real-time overlaps and intersections between factors using a biopsychosocial, whole-person perspective (Herbert et al., 2022). By capturing complex longitudinal information about intraindividual variation in pain sites in a single number, our metric of pain site variation is ideal to support such research questions.

## Data Availability

Data analysis script code is available on Open Science Framework. The de-identified data are available for selective sharing, subject to review of individual data requests, the use of secure computer platforms, formal use agreements, and compliance with the institutional human research ethics policies. Data use is limited to research purposes, on secure computer servers. The principal investigator (VJM) is the contact person for requests to share data.

https://osf.io/xuvjg/overview?view_only=4f4375b56f764d6c86c616744a313989

## Acknowledgements

We thank Nomvula Mdwaba, Ncumisa Msolo, and Andiswa Gidana for supporting data collection.

## Author contributions

This study was led by VJM. All authors contributed to conceptualisation, methodology, and the investigation. Formal analysis and the original writing were led by VJM, GA, and PK. All authors gave feedback on the results and their presentation, reviewed and edited the writing and have approved the final version of the manuscript to be submitted.

