## Supplementary file for "Quantifying intra-individual variations in anatomical sites of pain in longitudinal studies"

### Supplementary material

#### Methods

##### Outcomes

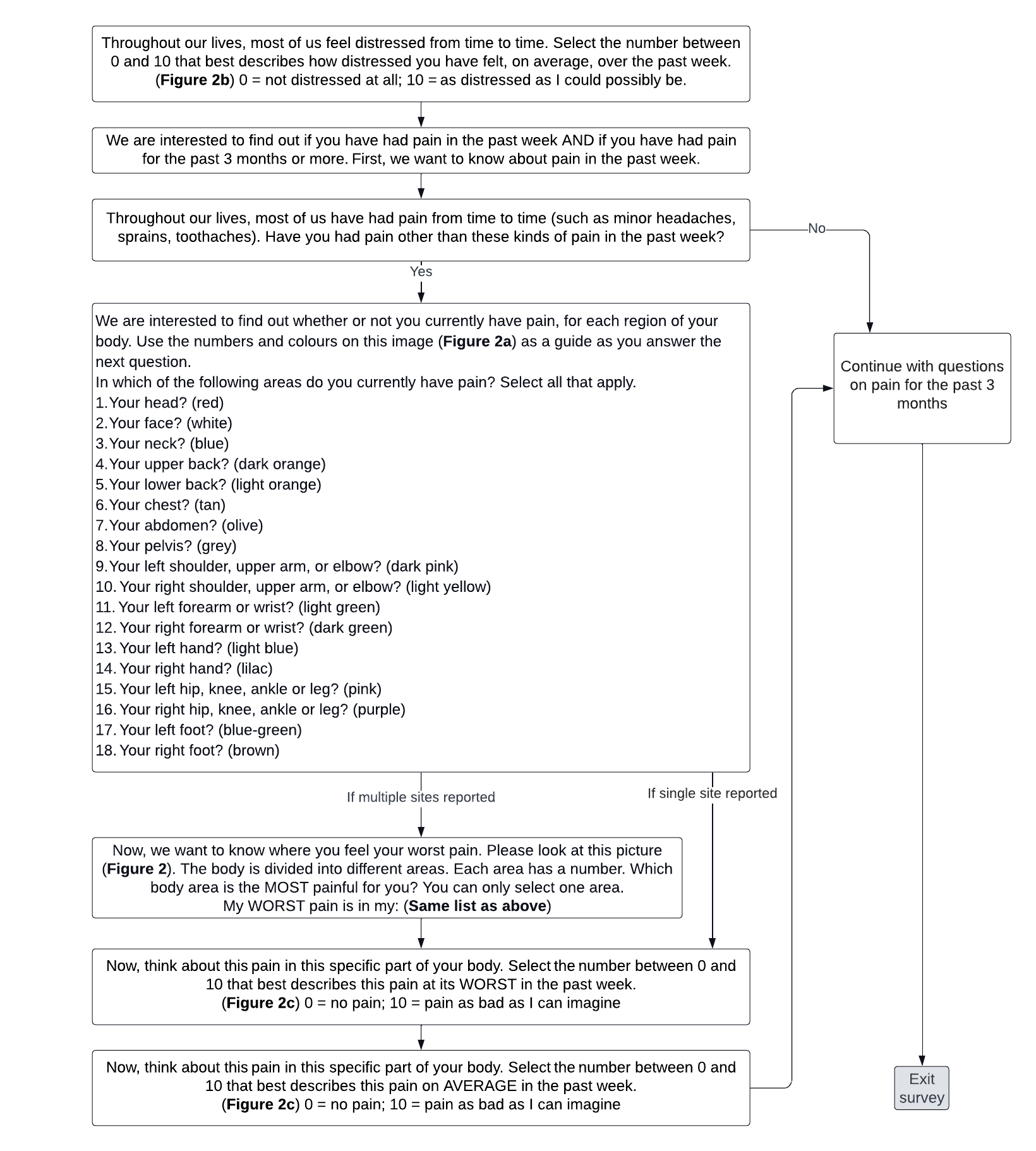

Figure S1: Flow chart with complete questions.

#### **Results**

#### Description of all data

*Table S1: Unadjusted linear regression models for number of missing responses as outcome, with age, sex, mean distress rating, mean of the worst pain severity and the mean of the count of painful sites as predictors*

|  | **Age model** | | | **Sex model** | | | **Distress model** | | | **Pain intensity model** | | | **Count of painful sites model** | | |
| --- | --- | --- | --- | --- | --- | --- | --- | --- | --- | --- | --- | --- | --- | --- | --- |
| *Predictors* | *Estimates* | *CI* | *p* | *Estimates* | *CI* | *p* | *Estimates* | *CI* | *p* | *Estimates* | *CI* | *p* | *Estimates* | *CI* | *p* |
| Intercept | 25.61 | 14.31 – 36.91 | **<0.001** | 32.02 | 29.58 – 34.46 | **<0.001** | 37.13 | 33.69 – 40.57 | **<0.001** | 37.51 | 32.22 – 42.80 | **<0.001** | 30.81 | 27.35 – 34.26 | **<0.001** |
| Age | 0.19 | -0.06 – 0.45 | 0.130 |  |  |  |  |  |  |  |  |  |  |  |  |
| Sex (male) |  |  |  | 7.27 | 2.75 – 11.79 | **0.002** |  |  |  |  |  |  |  |  |  |
| Mean distress |  |  |  |  |  |  | -0.73 | -1.39 – -0.07 | **0.030** |  |  |  |  |  |  |
| Mean worst pain severity |  |  |  |  |  |  |  |  |  | -1.03 | -1.78 – -0.27 | **0.009** |  |  |  |
| Mean count of painful sites |  |  |  |  |  |  |  |  |  |  |  |  | 0.43 | -0.53 – 1.39 | 0.375 |
| Observations | 72 | | | 72 | | | 72 | | | 52 | | | 56 | | |
| R^2^ / R^2^ adjusted | 0.033 / 0.019 | | | 0.128 / 0.116 | | | 0.065 / 0.052 | | | 0.130 / 0.113 | | | 0.015 / -0.004 | | |

##### Pain sites variation metric

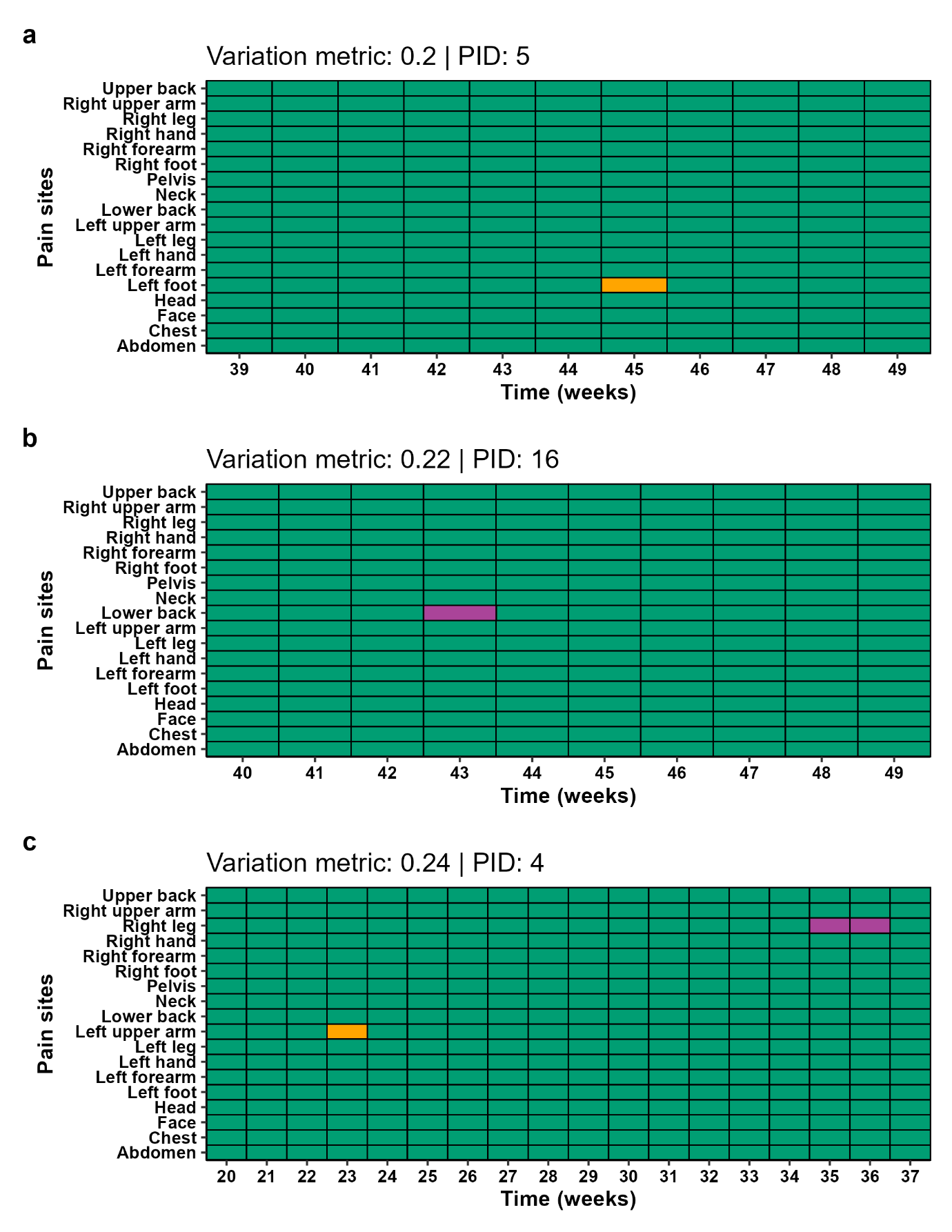

Figure S2: Pain sites (in grey) and the worst pain site (in dark grey) endorsed by three participants (a-c) with the lowest variation metric (0.20-0.24).

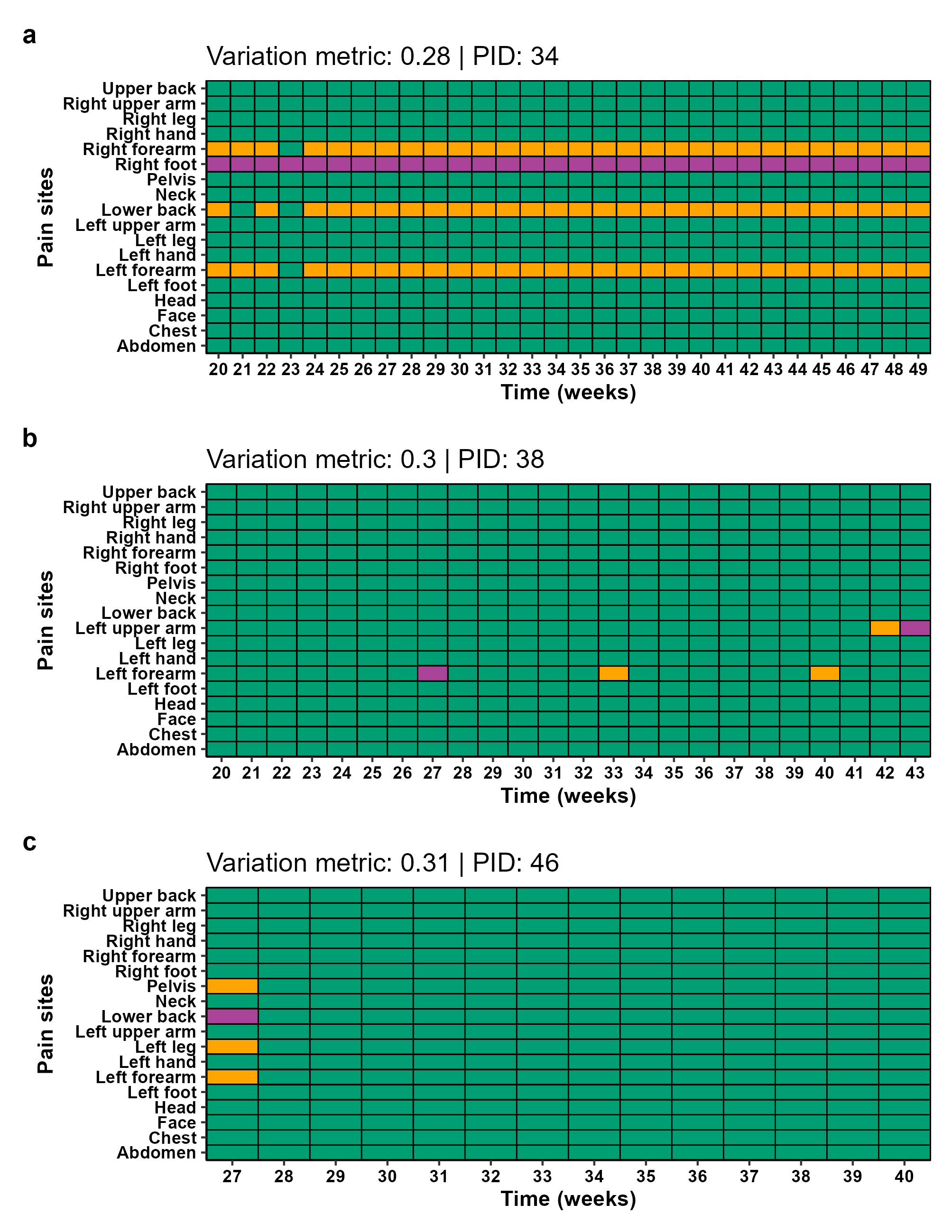

Figure S3: Pain sites (in grey) and the worst pain site (in dark grey) endorsed by three participants (a-c) with a variation metric between 0.28 and 0.31.

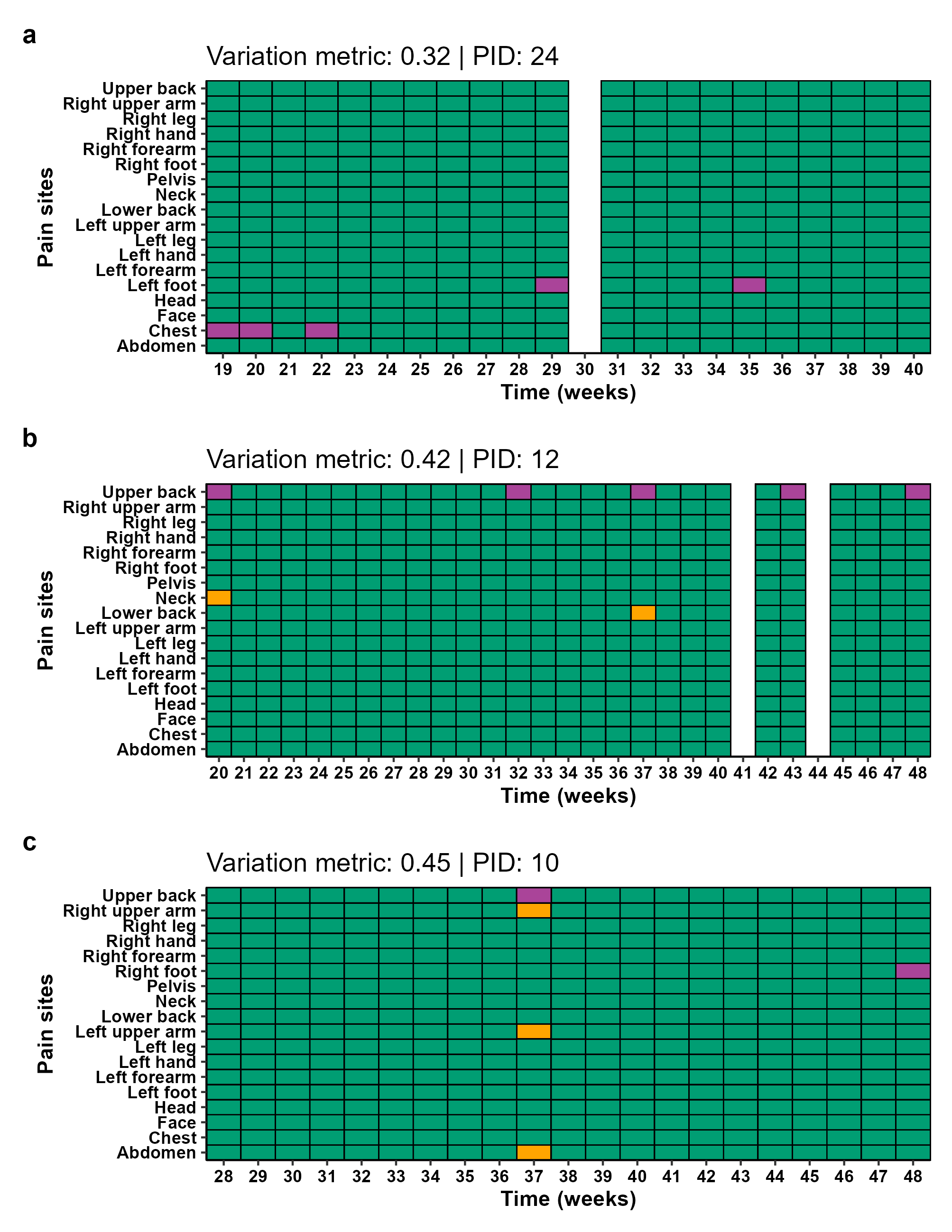

Figure S4: Pain sites (in grey) and the worst pain site (in dark grey) endorsed by three participants (a-c) with a variation metric between 0.32 and 0.45. White shows participant not responding that week.

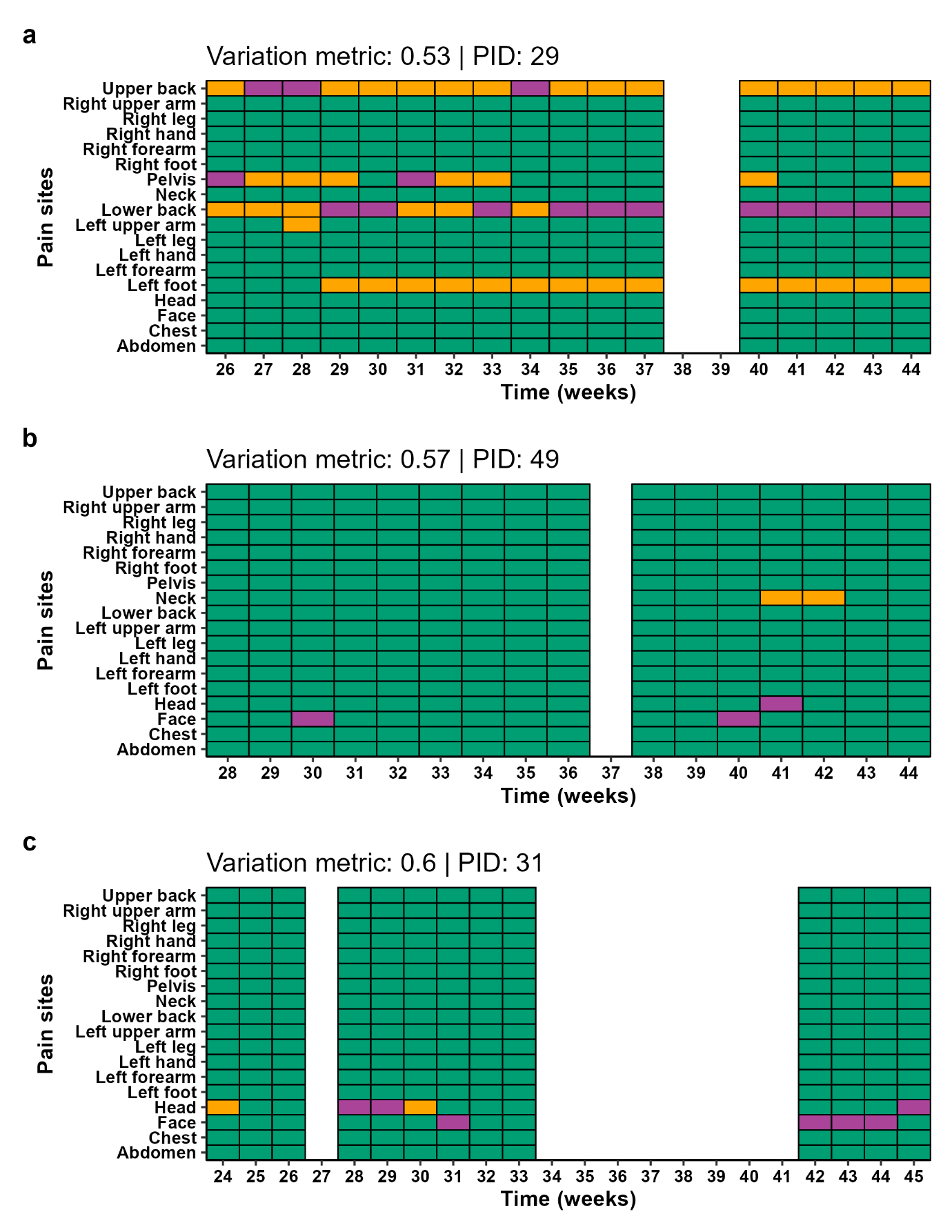

Figure S5: Pain sites (in grey) and the worst pain site (in dark grey) endorsed by three participants (a-c) with a variation metric between 0.53 and 0.60. White shows participant not responding that week.

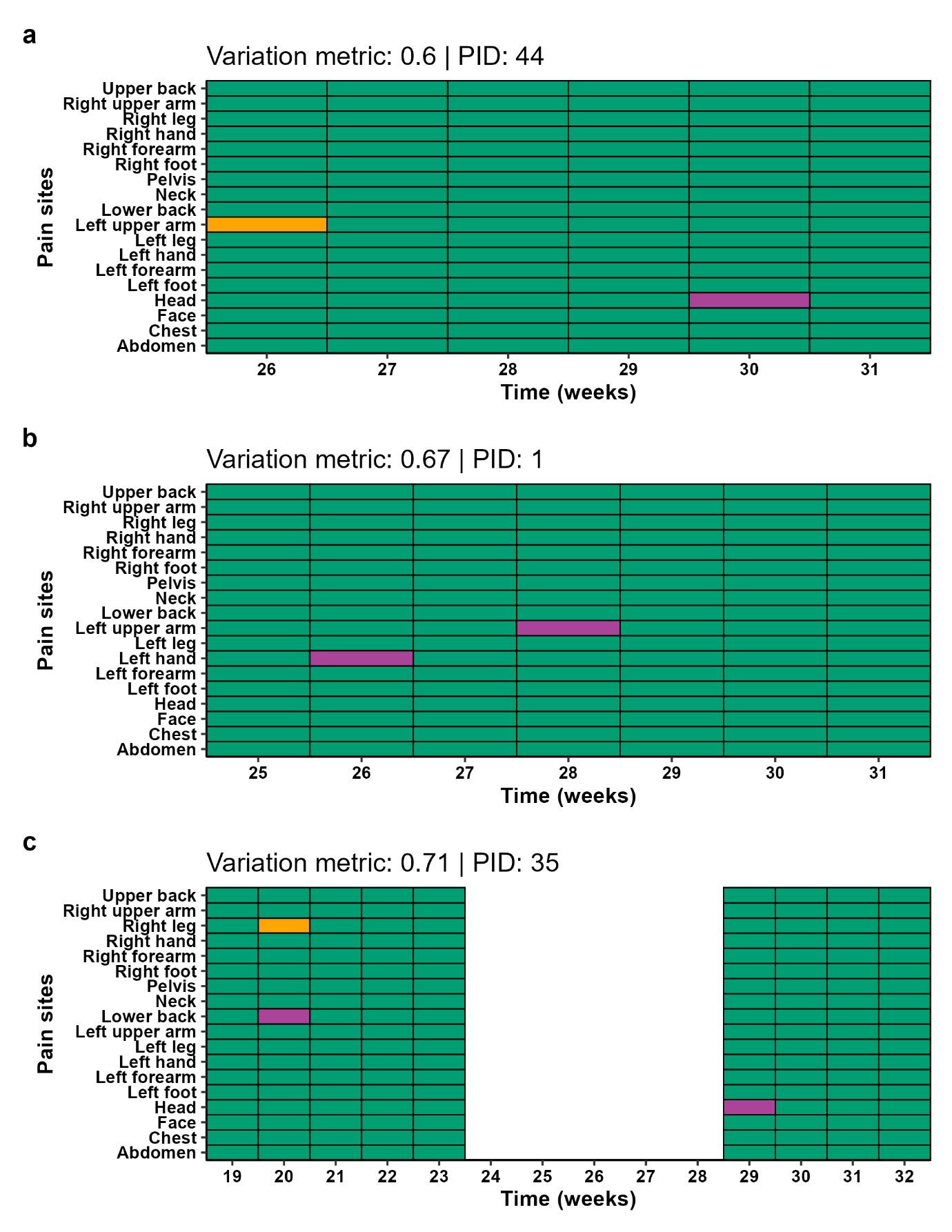

Figure S6: Pain sites (in grey) and the worst pain site (in dark grey) endorsed by three participants (a-c) with a variation metric between 0.60 and 0.71. White shows participant not responding that week.

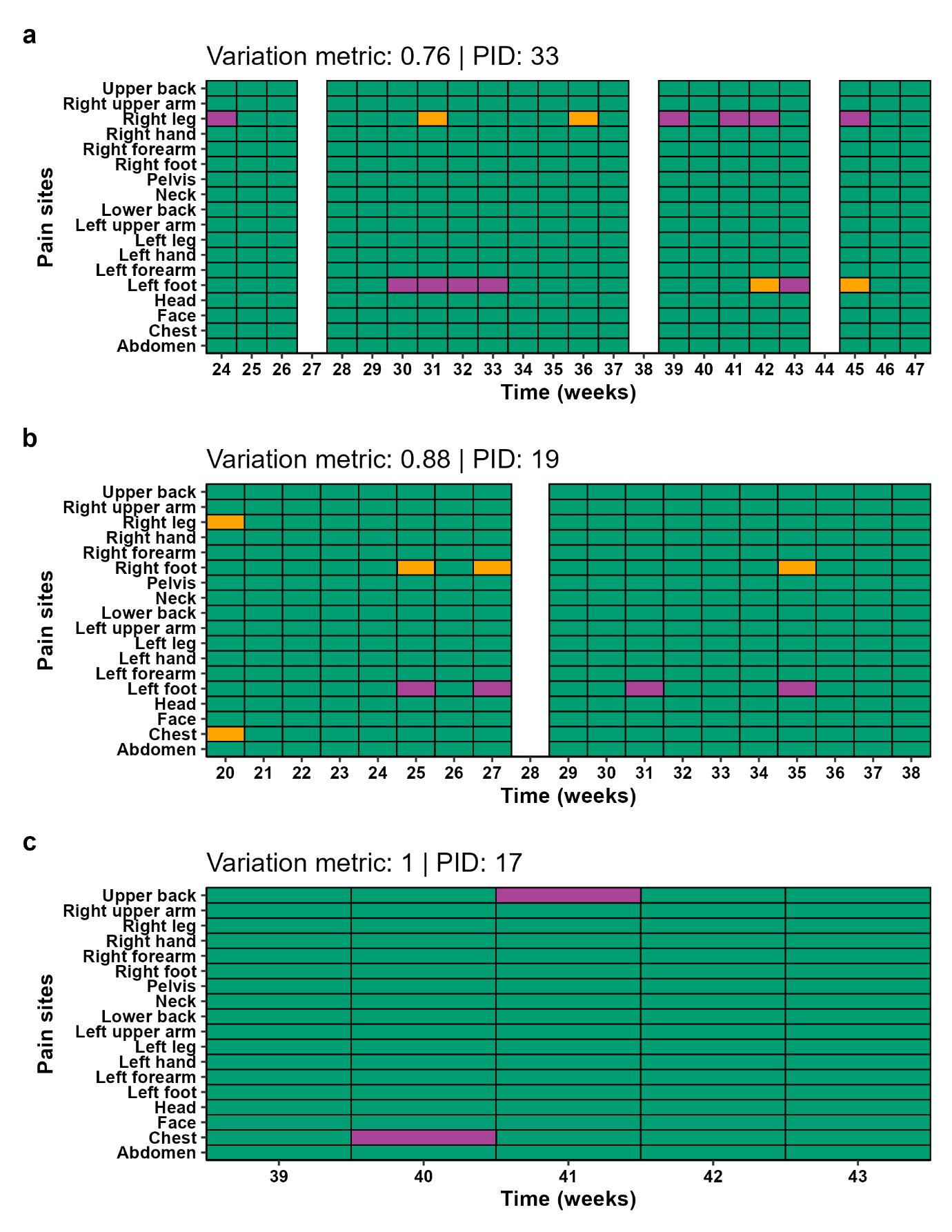

Figure S7: Pain sites (in grey) and the worst pain site (in dark grey) endorsed by three participants (a-c) with a variation metric between 0.76 and 1.00. White shows participant not responding that week.

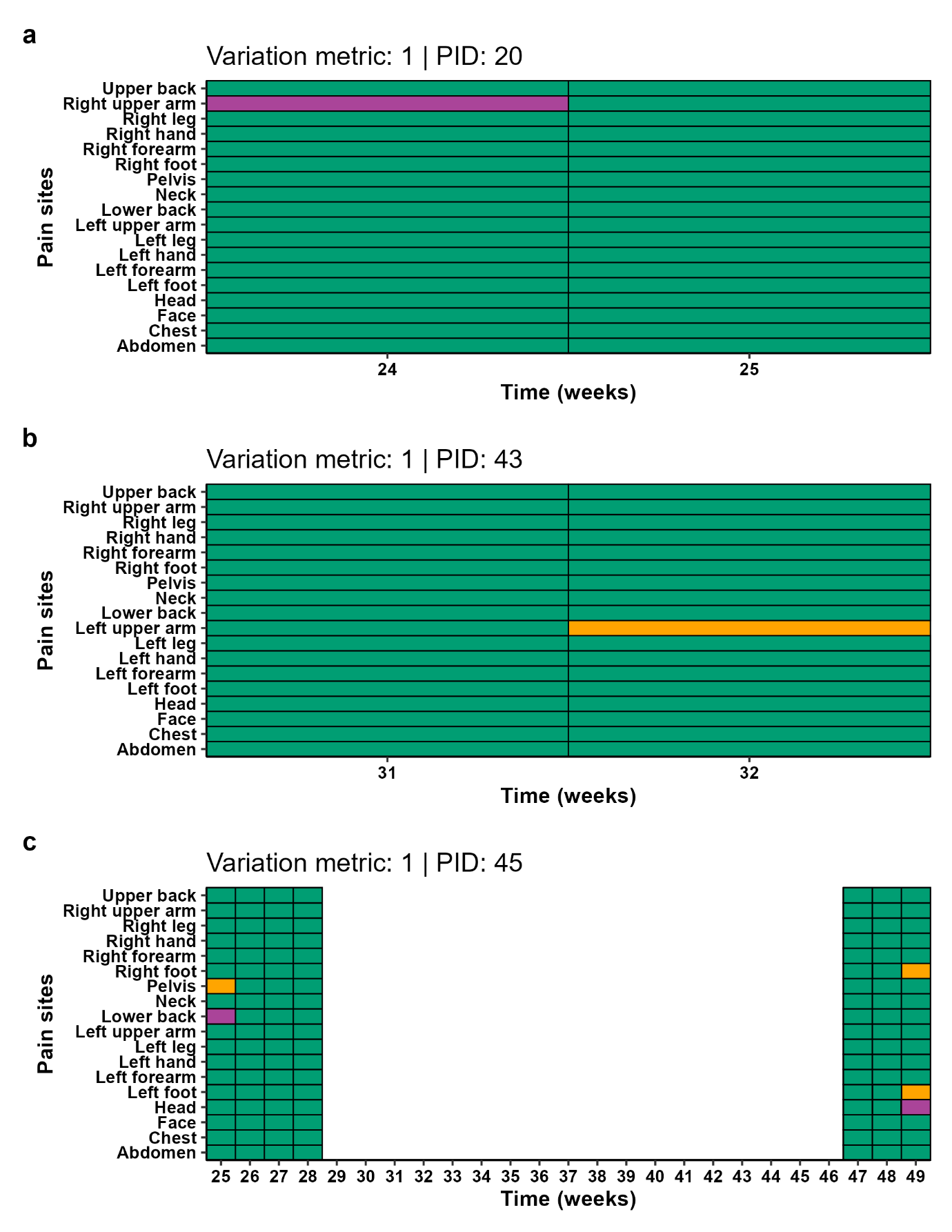

Figure S8: Pain sites (in grey) and the worst pain site (in dark grey) endorsed by three participants (a-c) with a variation metric less of 1.00. White shows participant not responding that week.

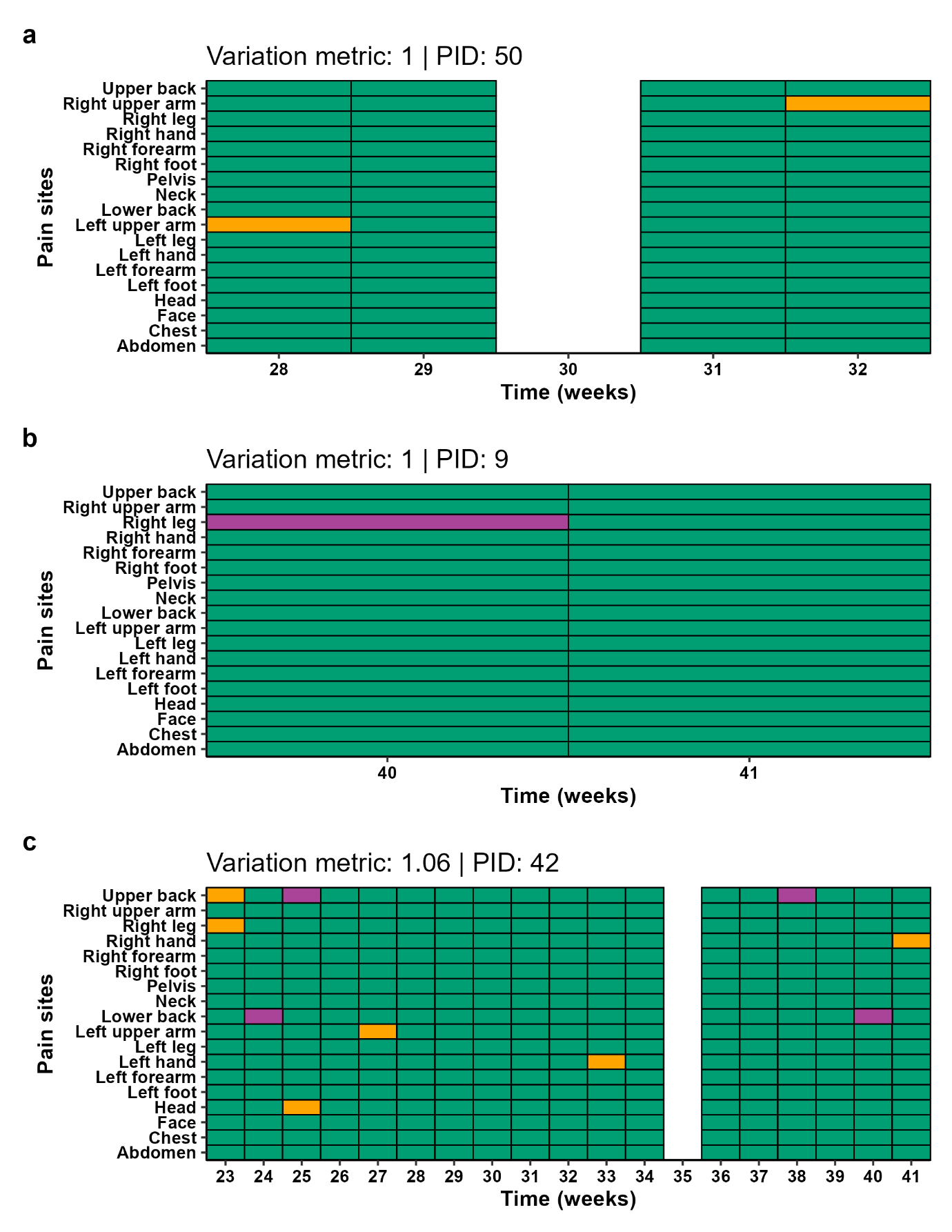

Figure S9: Pain sites (in grey) and the worst pain site (in dark grey) endorsed by three participants (a-c) with a variation metric less between 1.00 and 1.06. White shows participant not responding that week.

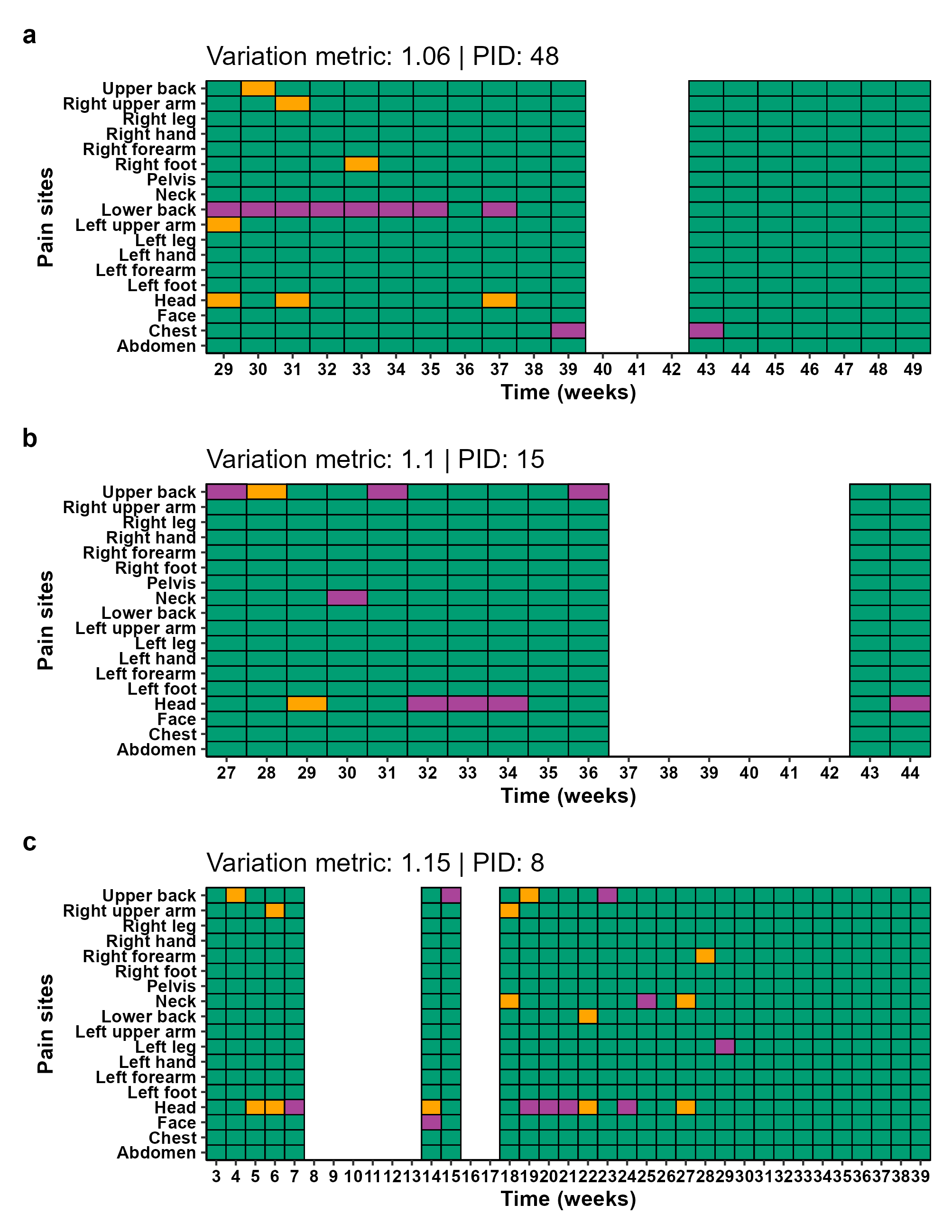

Figure S10: Pain sites (in grey) and the worst pain site (in dark grey) endorsed by three participants (a-c) with a variation metric less between 1.06 and 1.15. White shows participant not responding that week.

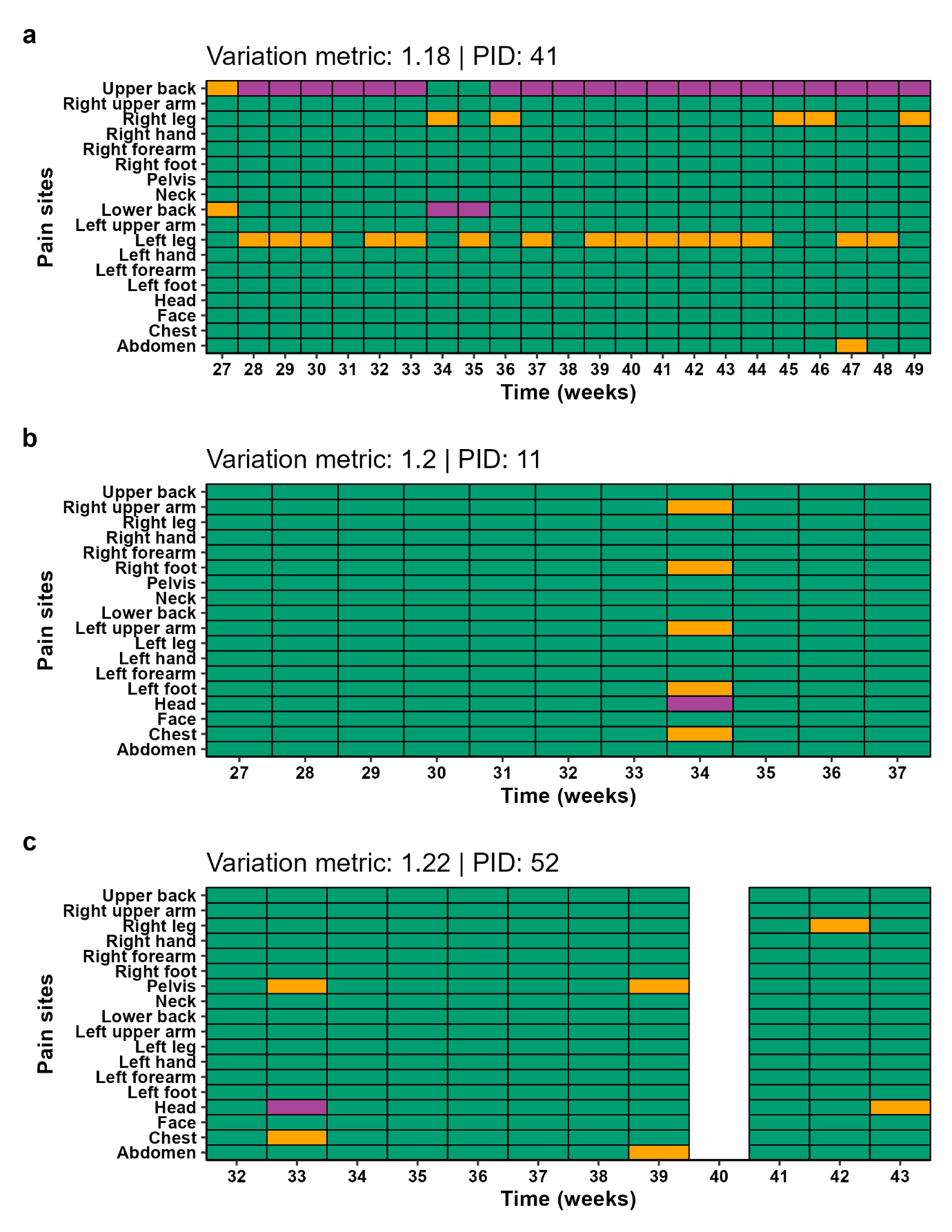

Figure S11: Pain sites (in grey) and the worst pain site (in dark grey) endorsed by three participants (a-c) with a variation metric between 1.18 and 1.22. White shows participant not responding that week.

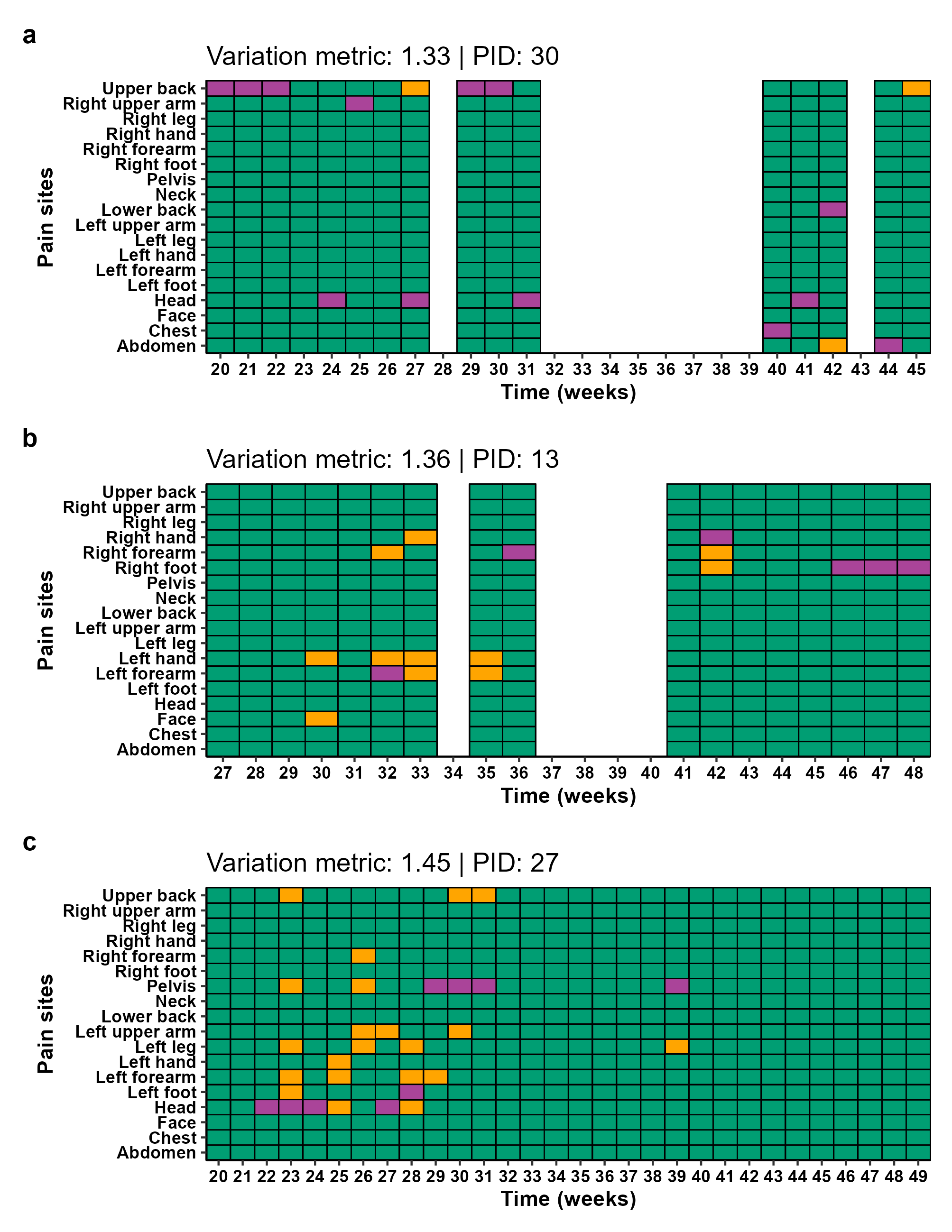

Figure S12: Pain sites (in grey) and the worst pain site (in dark grey) endorsed by three participants (a-c) with a variation metric between 1.33 and 1.45. White shows participant not responding that week.

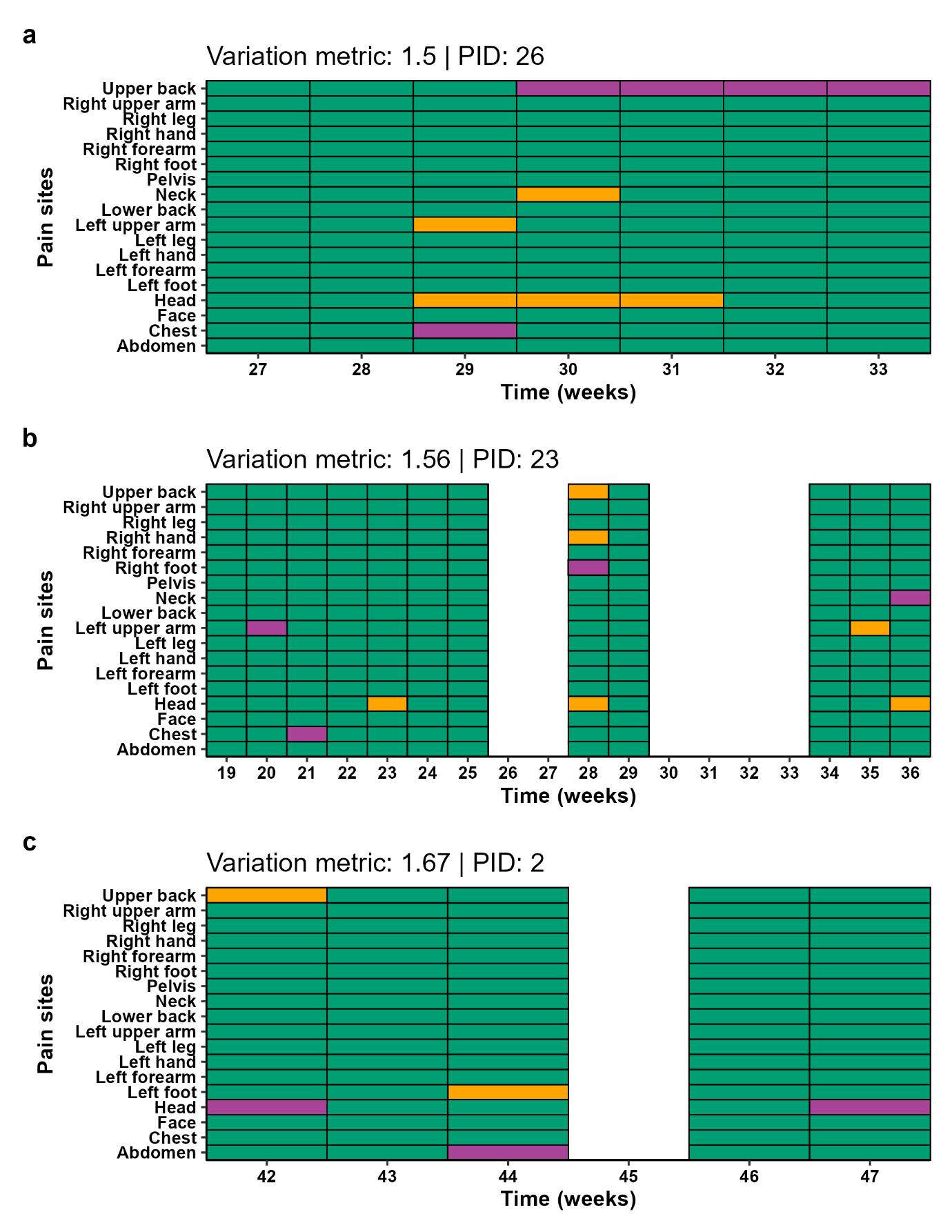

Figure S13: Pain sites (in grey) and the worst pain site (in dark grey) endorsed by three participants (a-c) with a variation metric between 1.50 and 1.67. White shows participant not responding that week.

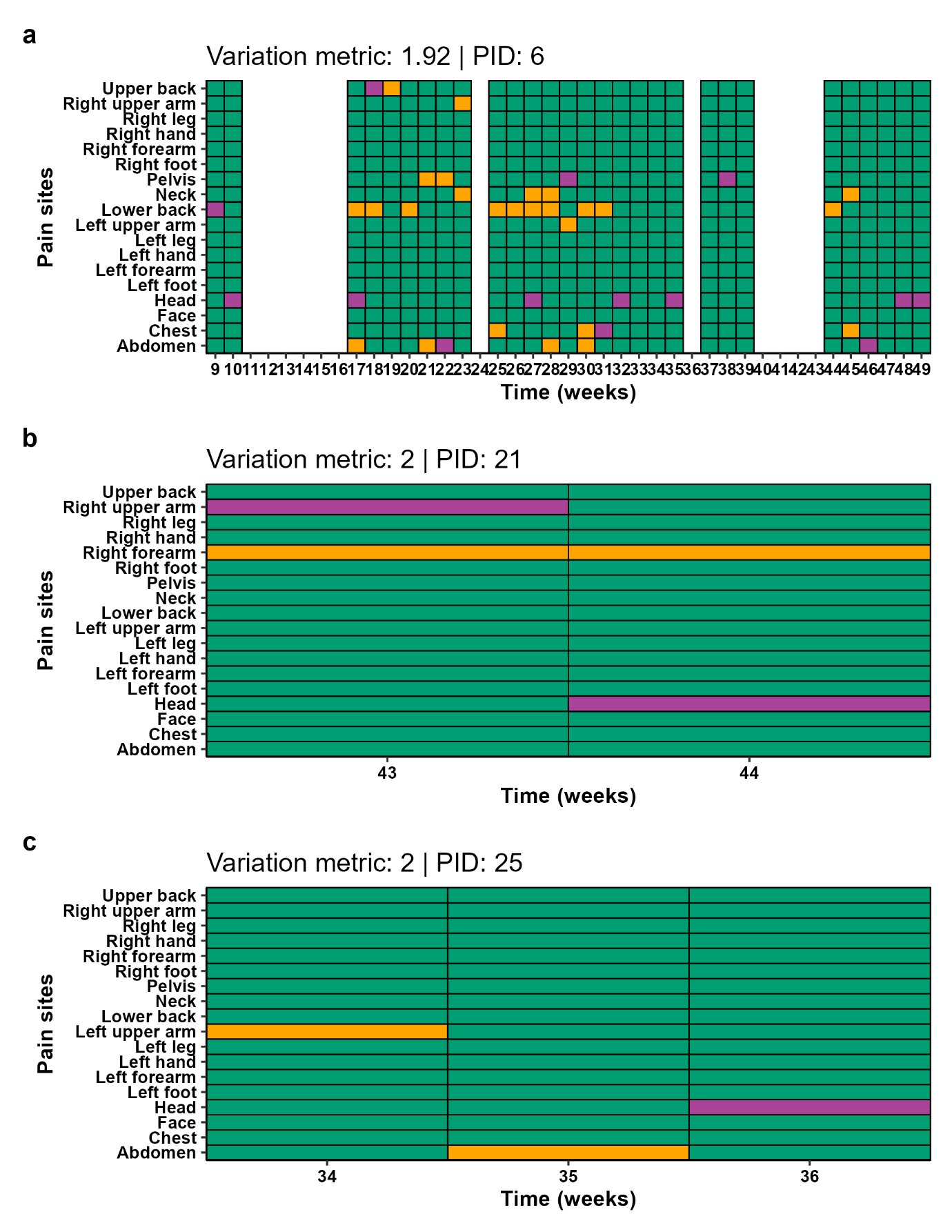

Figure S14: Pain sites (in grey) and the worst pain site (in dark grey) endorsed by three participants (a-c) with a variation metric between 1.92 and 2.00. White shows participant not responding that week.

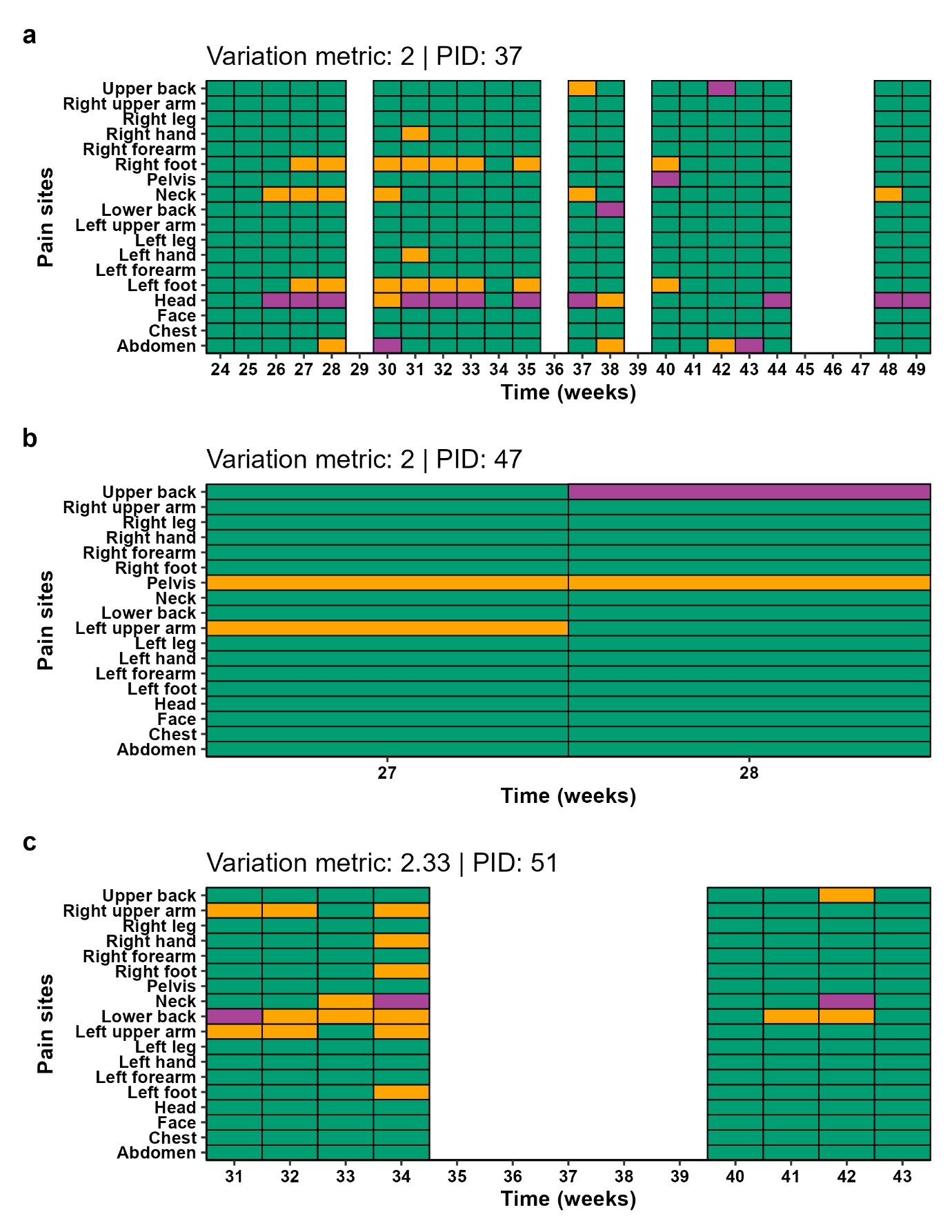

Figure S15: Pain sites (in grey) and the worst pain site (in dark grey) endorsed by three participants (a-c) with a variation metric between 2.00 and 2.33. White shows participant not responding that week.

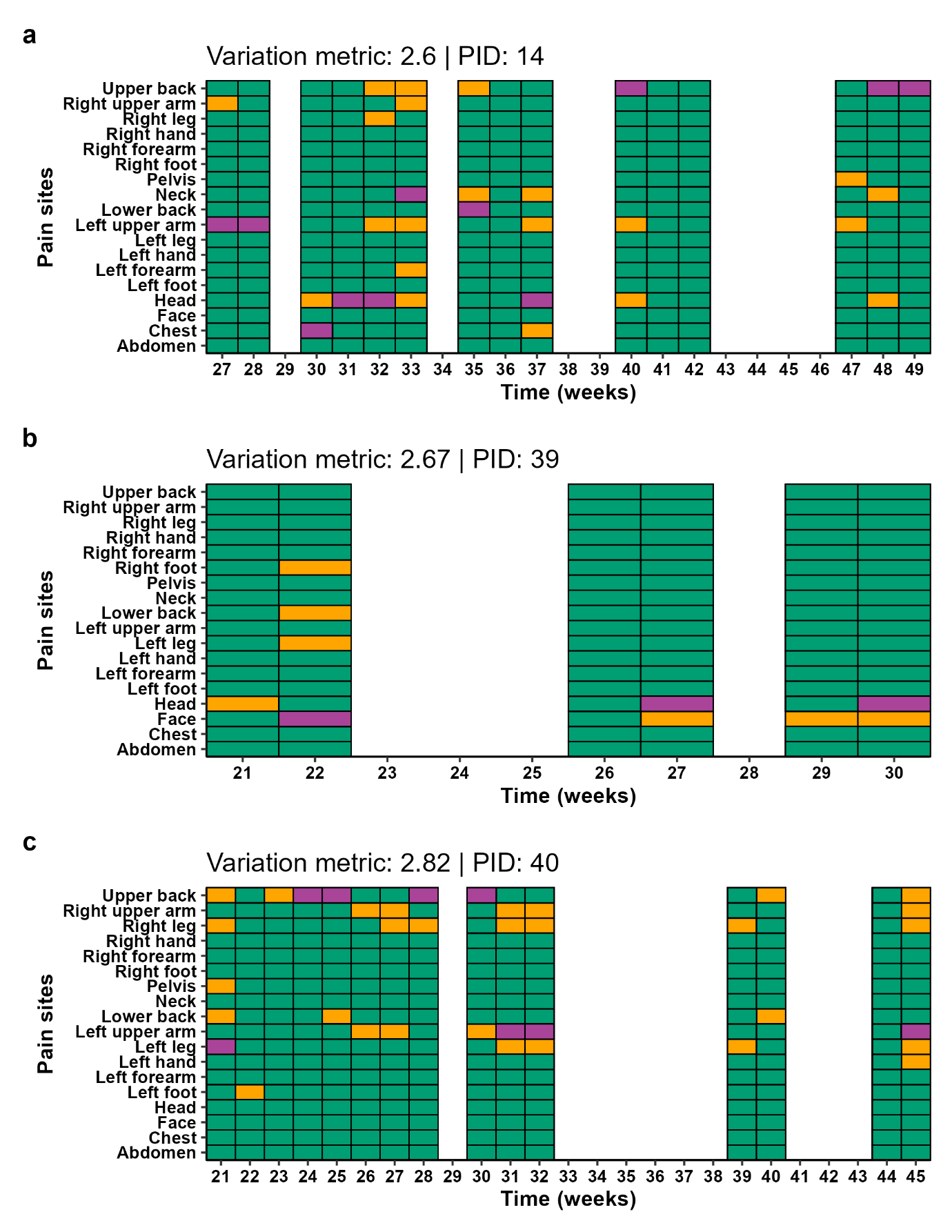

Figure S16: Pain sites (in grey) and the worst pain site (in dark grey) endorsed by three participants (a-c) with a variation metric between 2.60 and 2.82. White shows participant not responding that week.

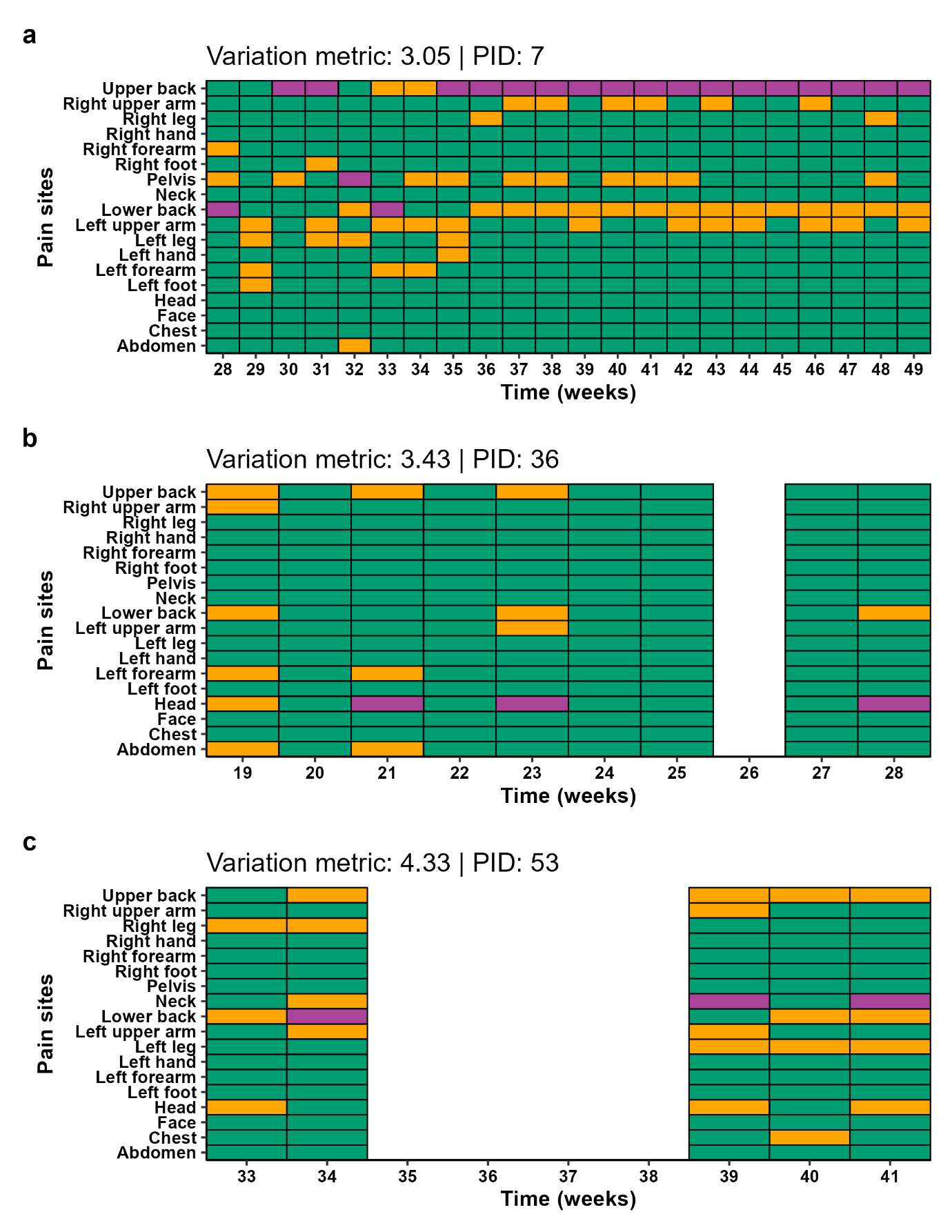

Figure S17: Pain sites (in grey) and the worst pain site (in dark grey) endorsed by three participants (a-c) with a variation metric between 3.05 and 4.33. White shows participant not responding that week.

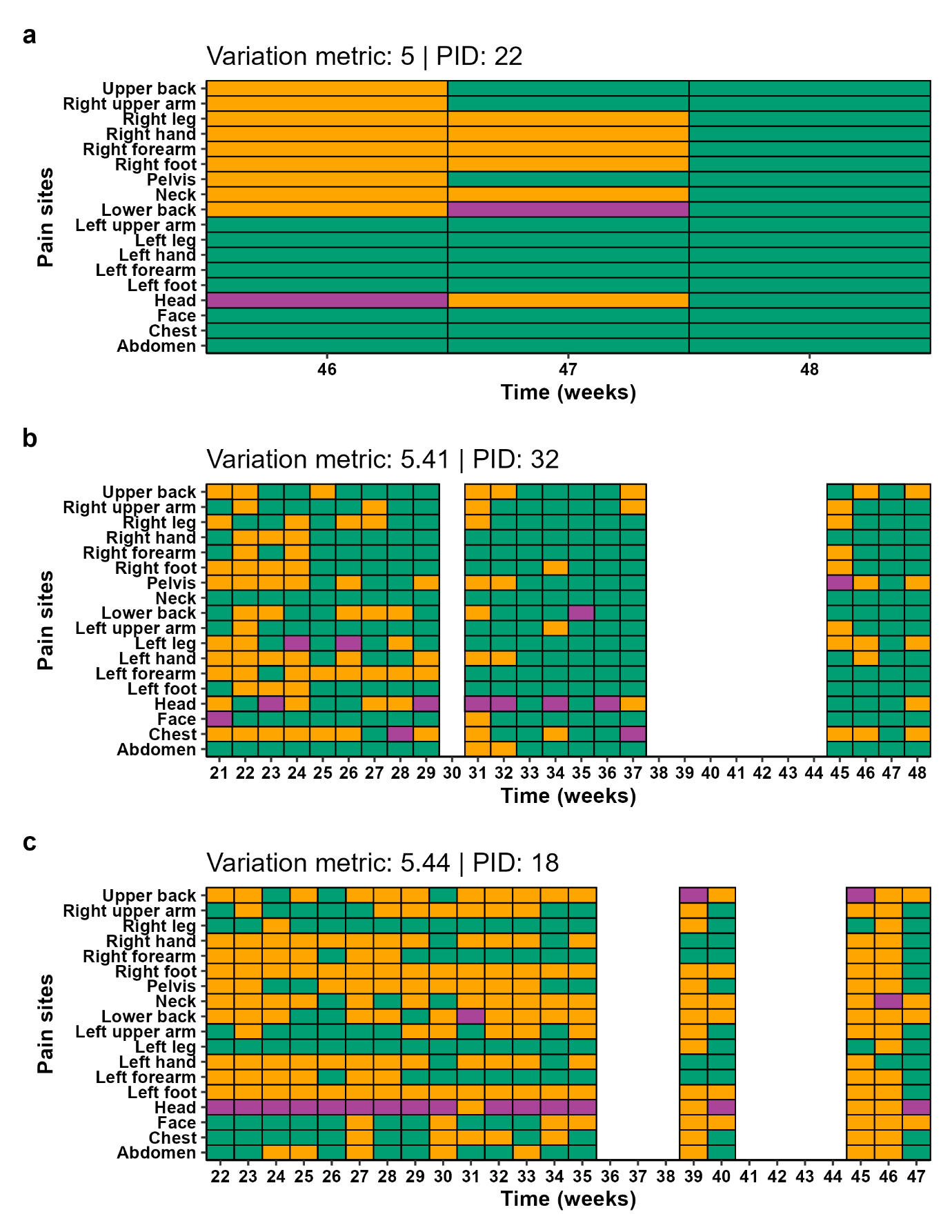

Figure S18: Pain sites (in grey) and the worst pain site (in dark grey) endorsed by three participants (a-c) with a variation metric between 5.00 and 5.44. White shows participant not responding that week.

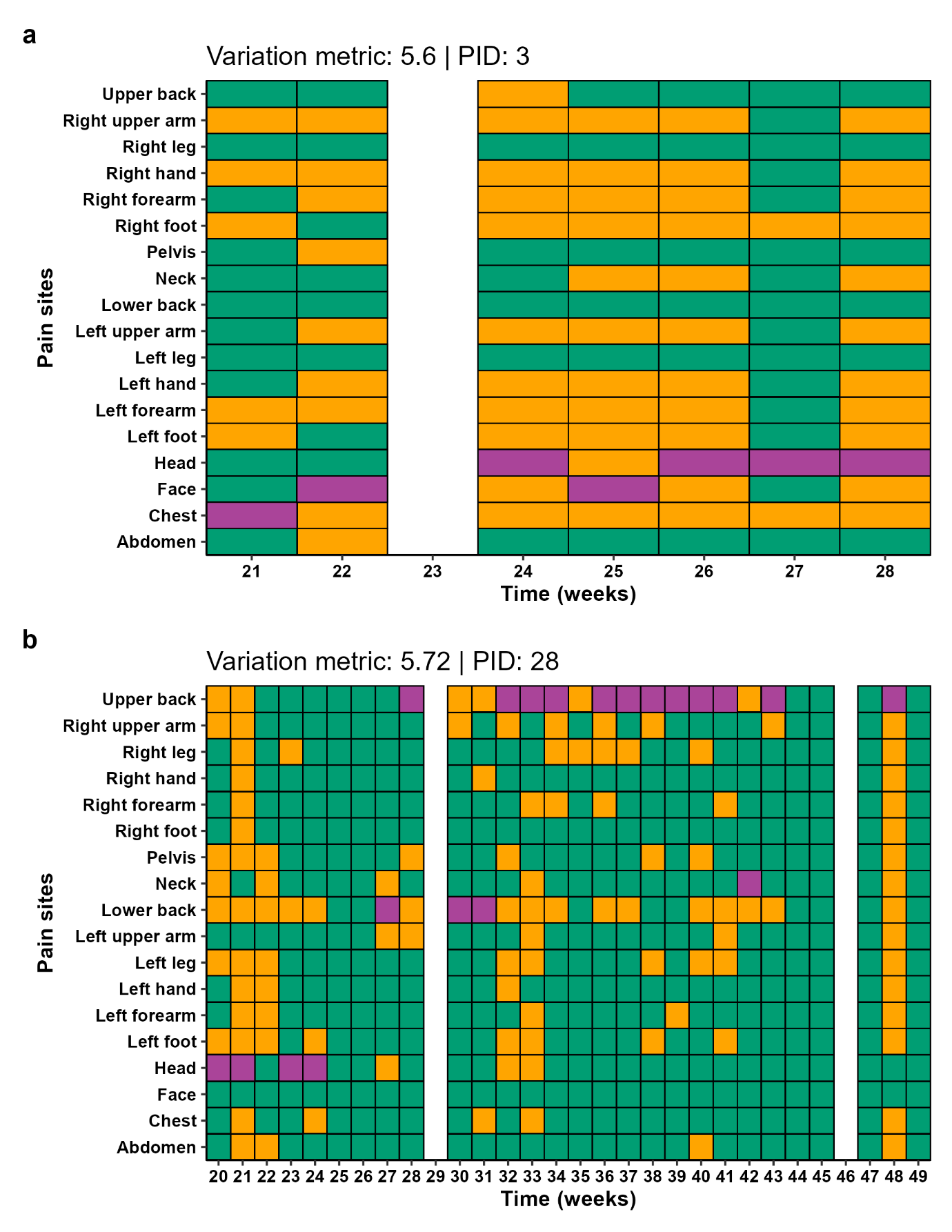
Figure S19: Pain sites (in grey) and the worst pain site (in dark grey) endorsed by two participants (a & b) with the highest variation metric (5.60 & 5.72).

###### Correlation analysis

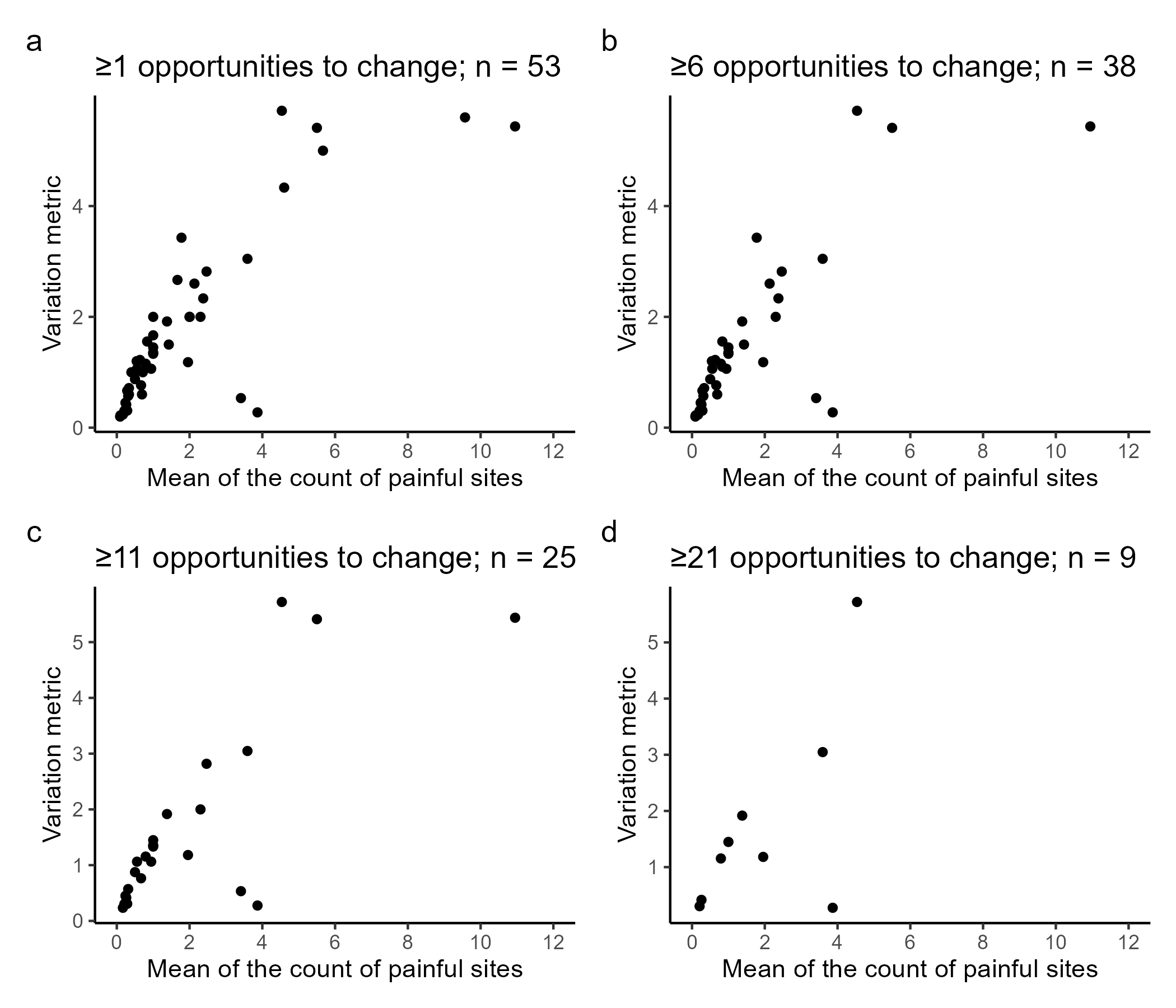

Figure S20: Relationships between the pain site variation metric and the mean of count of painful sites, at cutoffs for participant inclusion (a-d).

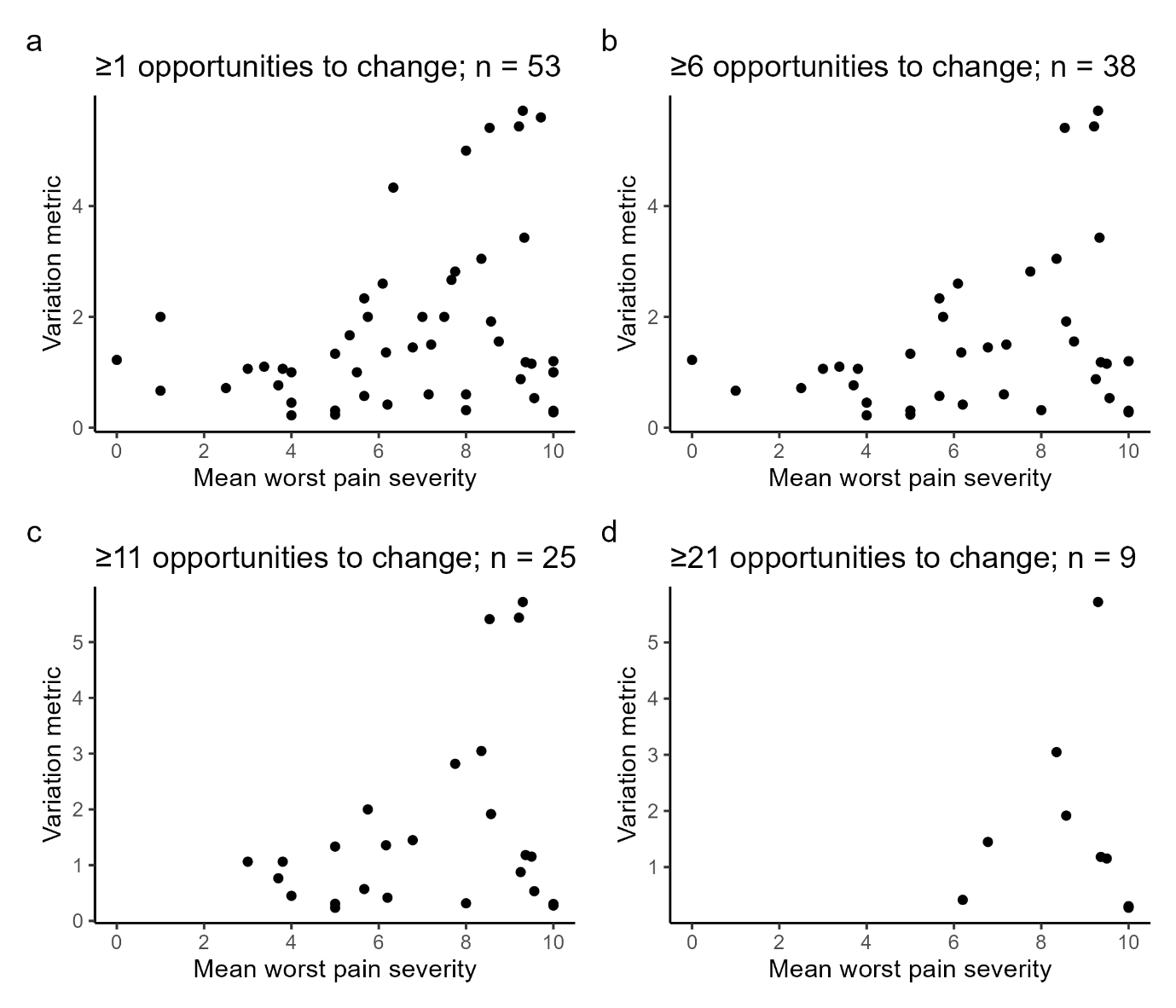

Figure S21: Relationships between the pain site variation metric and the mean worst pain, at cutoffs for participant inclusion (a-d).

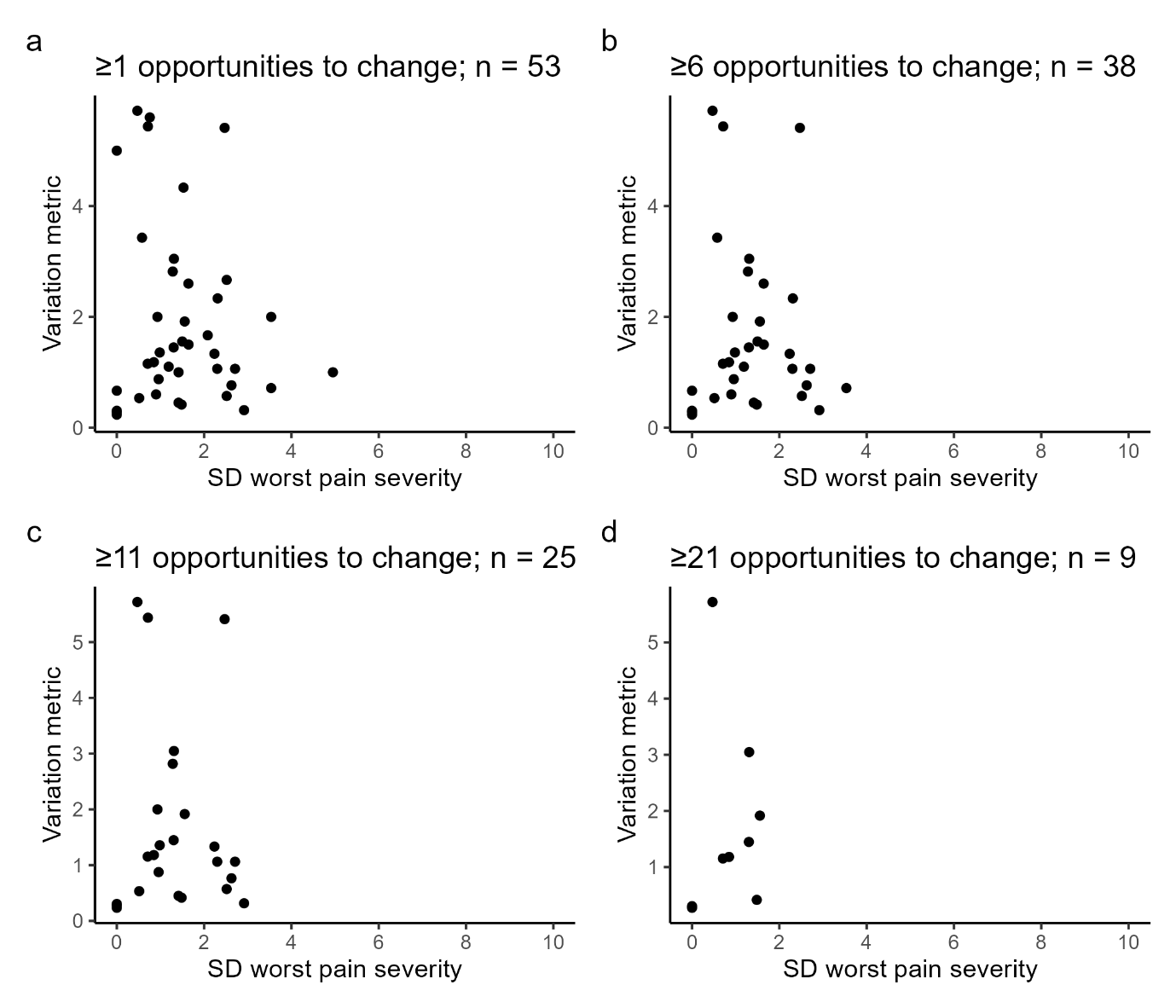

Figure S22: Relationships between the pain site variation metric and the standard deviation of worst pain, at cutoffs for participant inclusion (a-d).

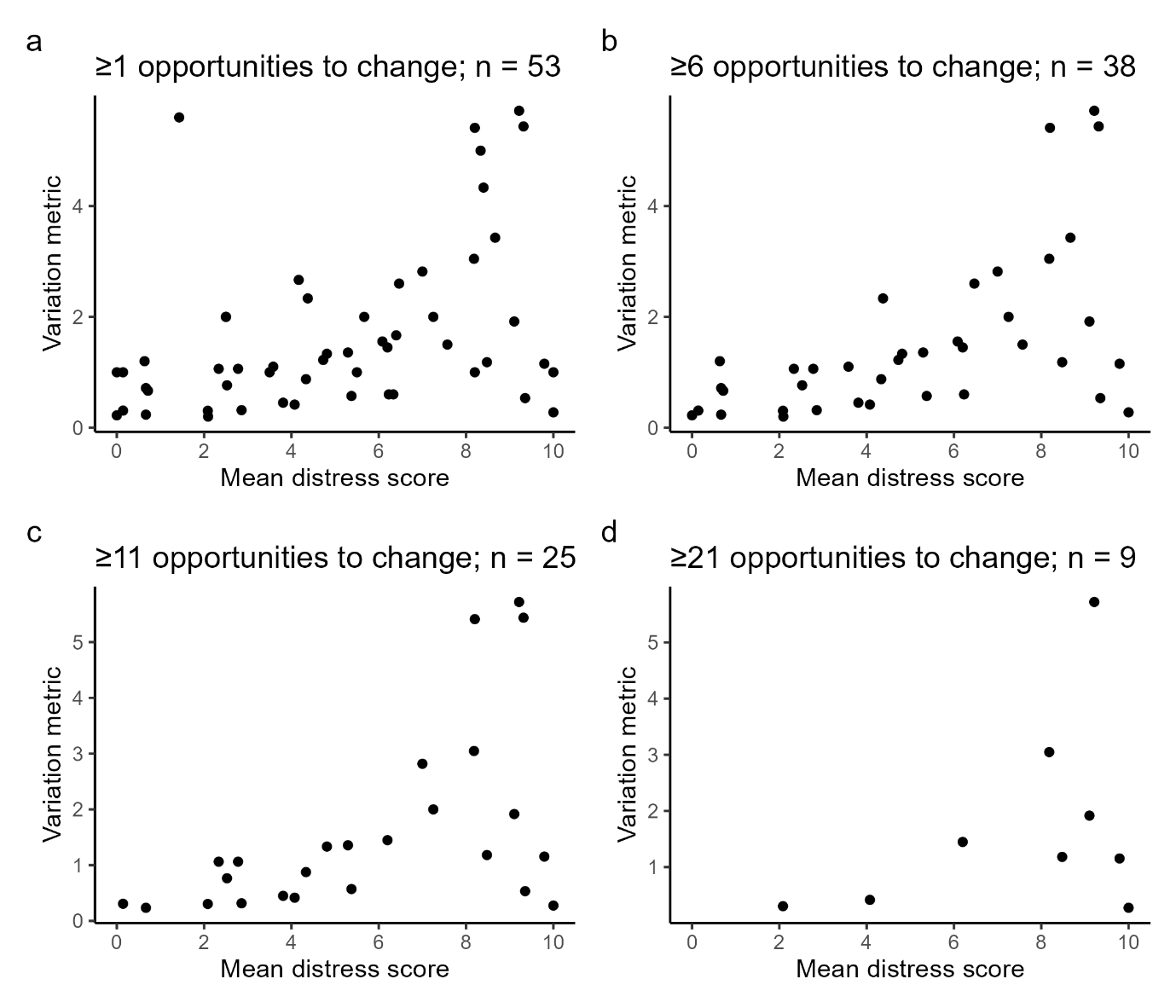

Figure S23: Correlations between the pain site variation metric and mean distress, with different cutoffs for participant inclusion (a-d).
